# Digital Adherence Support for Tuberculosis Treatment: A Multicentre Randomized Trial in Kenya

**DOI:** 10.64898/2026.02.11.26346015

**Authors:** Erez Yoeli, Jonathan Rathauser, Edwin Nyakan, Justin J. Boutilier, Jonathon R. Campbell, Moreen Chilo, Lewis Muriuki Irungu, Jónas Oddur Jónasson, Maureen Kamene Kimenye, Faith Kavata Muchiri, Alice M. Mwikamba, Teresa Adhiambo Ochieng, Jill Ondigo, Philip Owiti, Kevin Schwartzman, David G. Rand

## Abstract

**Background:** Improving tuberculosis (TB) treatment success is critical for improving the health of individuals with TB, reducing transmission, and lowering treatment costs. We conducted a four-arm randomized controlled trial (RCT) to evaluate whether three digital interventions with increasing support improved treatment outcomes compared to the standard of care.

**Methods:** In this open-label, parallel RCT in Kenya, all TB patients at 902 participating clinics who had at least 2 months of treatment remaining were eligible for inclusion. Individuals were centrally randomized at a ratio of 4:3:12:12 into a control group that received only the standard of care, or one of the following three intervention groups that received the standard of care plus: (1) a daily SMS medication adherence reminder (‘SMS’); (2) access to a platform that sent a daily request for self-verification of medication adherence and provided disease information, motivational messages, and an adherence game (‘platform’); and (3) access to a platform with these same features, plus support from a team of trained supporters (‘Keheala’). The primary outcomes were: the proportion of individuals who experienced an unsuccessful treatment outcome (a composite of: died, failed treatment, or loss to follow-up), and loss to follow-up (LTFU). The secondary outcome was medication non-adherence, measured via unannounced urine isoniazid tests for a random sample of 731 individuals in the control and Keheala groups.

**Results:** Between April 13, 2018 and December 20, 2019, 16,753 individuals were randomized, yielding 14,962 in the mITT population: 1,997 in the control, 1,475 in SMS, 6,057 in platform, and 5,433 in Keheala. Absolute risk of unsuccessful outcomes was 12.4% in the control group. It was reduced by 1.9 percentage points in the SMS group (95% C.I.: -0.1–4.0), 1.9 percentage points in the platform group (95% C.I.: 0.3–3.4), and 2.6 percentage points in the Keheala group (95% C.I.: 1.0-4.2); mostly due to reductions in LTFU. Medication non-adherence was 12.4% in the control group. It was reduced by 7.5 percentage points in the Keheala group (95% C.I.: 2.6-12.5).

**Conclusions:** All digital health interventions improved treatment outcomes. The Keheala intervention also reduced medication non-adherence. The interventions could be considered as a supplement to the standard of care, especially in resource-constrained regions where in-person support is impractical.

## Introduction

Improving support for individuals treated for tuberculosis (TB) disease is a major priority for governments and development agencies [1]. Digital adherence support (DAS) has the potential to address shortfalls in the current standard of care [2]. However, there are relatively few large, representative RCTs studying the effects of DAS, particularly on TB treatment outcomes [3, 4, 5]. This has contributed to 2022 WHO TB Care and Support Guidelines identifying the evaluation of “the effectiveness of different forms of interventions to improve treatment adherence” as a top research priority [6]. Such data would support TB programs’ efforts to implement DAS, which are presently hampered by a lack of sufficient evidence of the impact of interventions on medication adherence and treatment completion [3, 4, 7].

We developed a feature-phone compatible DAS intervention called Keheala, using techniques from the behavioral sciences for promoting prosocial behaviors [8]. In a 1,104-patient randomized controlled trial (RCT) in Nairobi, Kenya, Keheala reduced the absolute risk of unsuccessful TB treatment outcomes–a composite of loss to follow-up (LTFU), treatment failure, and death–by roughly two-thirds relative to a control group that received the standard of care [9]. While these results are promising, they leave a number of important questions unanswered, such as Keheala’s ability to operate at scale, help individuals from a diversity of backgrounds, reduce deaths per se, and impact medication adherence. Moreover, it is important to understand whether less intensive, and thus less costly, interventions that omit various of Keheala’s components would still work–which is especially relevant for application in resource constrained settings.

In order to address these gaps, we performed a four-arm, large-scale RCT throughout Kenya that evaluated the Keheala intervention, two DAS interventions with incrementally reduced support intensity, and a control group receiving the standard of care. We compared the effectiveness of each intervention to the standard of care for preventing all unsuccessful TB treatment outcomes, and loss to follow-up specifically. Additionally, for the Keheala intervention: (1) sample sizes were sufficient to identify a 1% reduction in absolute risk of death relative to the control, and (2) reductions in medication non-adherence were evaluated via unannounced urine isoniazied tests for randomly selected individuals in the Keheala and control groups.

## Methods

### Setting

Our study was conducted in Kenya, which reported 94,534 cases of TB in 2018, 84,345 in 2019, and 72,943 in 2020, a population case notification rate of between 259 and 300 per 100,000 [10]. Kenya’s rate of treatment non-completion–a composite of LTFU, treatment failure, and death–was 15.6% in 2018, 14.7% in 2019, 15.0% in 2020, and 11.0% in 2023 [11]. Mobile phone penetration in Kenya is over 90% [12, 13, 14].

Our RCT was conducted in partnership with 902 clinics distributed across eight counties in each of Kenya’s eight regions (see Fig. 1). The counties were: Kakamega, Kiambu, Kisumu, Machakos, Mombasa, Nairobi, Turkana and Wajir. These counties were selected in consultation with the National TB program to represent their region, e.g., to have a mix of both rural and urban clinics, and a mix of patient characteristics typical of the county as a whole. Once these counties were selected, all clinics within these counties were included in the RCT.

**Figure 1:**
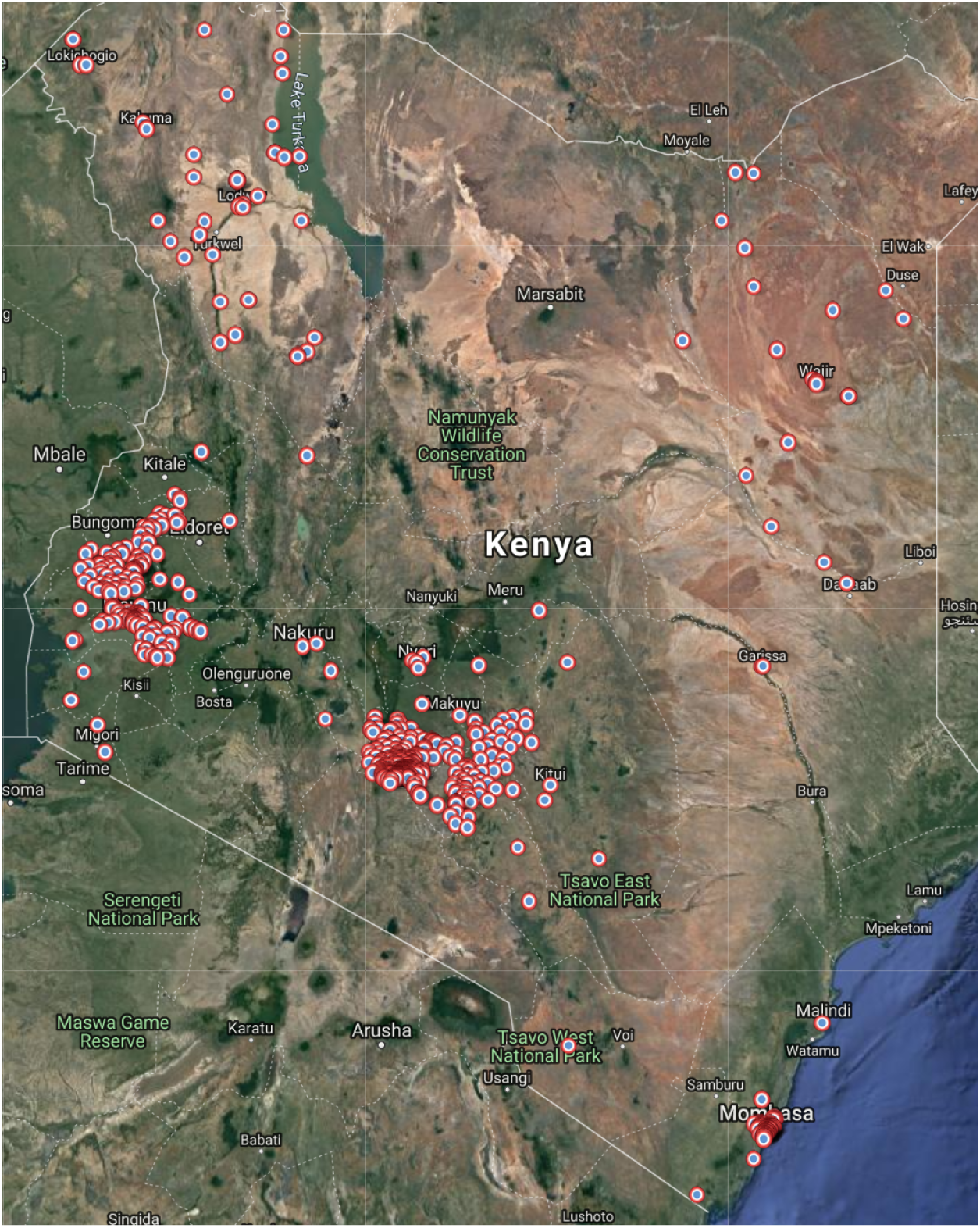
Partner Clinic Sites. Satellite map of Kenya overlaid with the location of the study’s 902 partner clinics. The clinics are located within the counties of Kakamega, Kiambu, Kisumu, Machakos, Mombasa, Nairobi, Turkana and Wajir.

**Figure 2:**
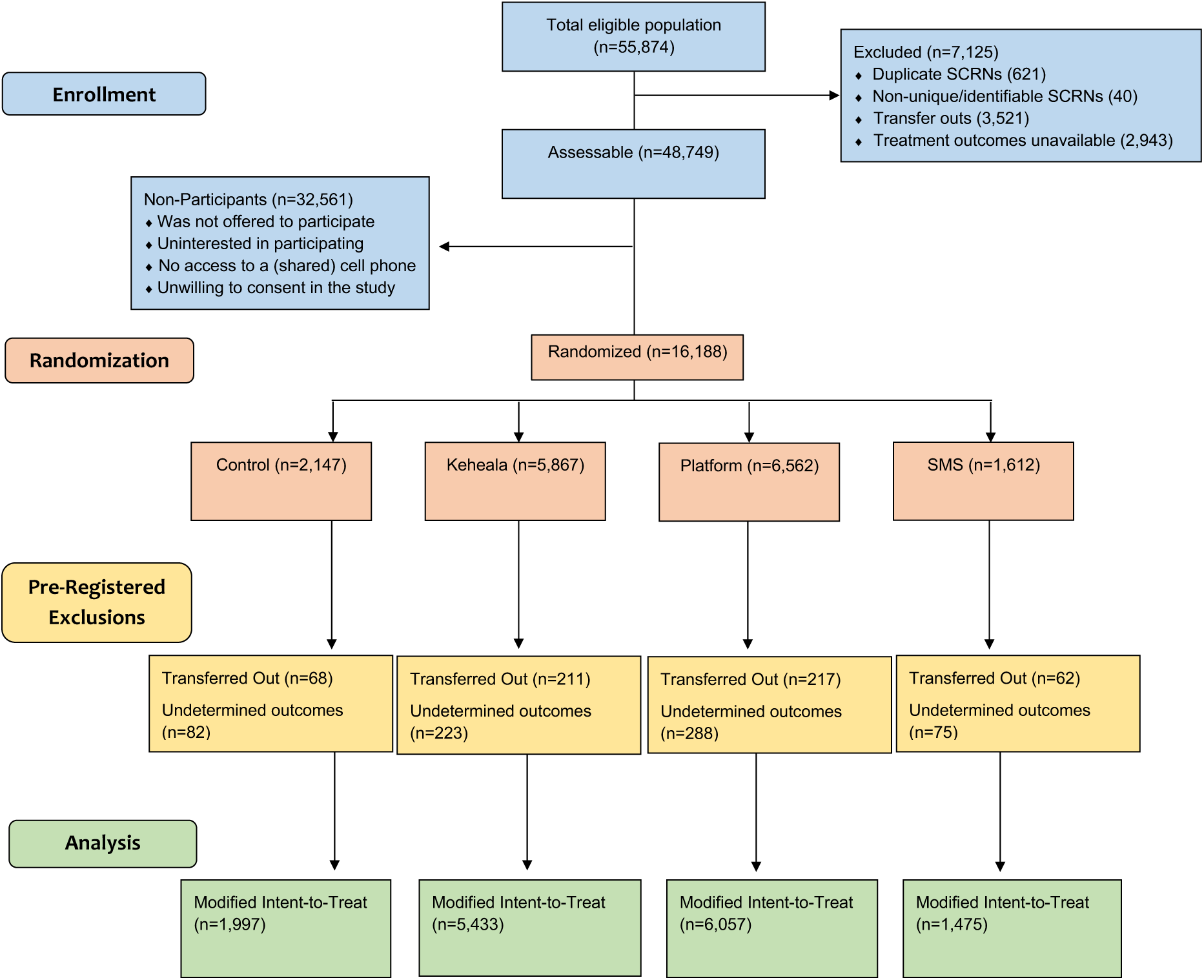
CONSORT Flow Diagram.

In Kenya, national guidelines call for directly observed treatment, but, in practice, medication adherence is rarely directly monitored. Individuals return to the clinic regularly–weekly for the first two months, then bi-weekly thereafter–to pick up the following week’s supply of medication. Often, visits include additional counseling [15]. Most patients receive six months of treatment, though this may be extended for drug-resistance or extrapulmonary disease [16].

### Study Population

Individuals receiving treatment for TB at participating clinics were eligible for the study if they: (1) had been clinically diagnosed or bacteriologically confirmed to have TB; (2) could communicate in either Kiswahili or English; (3) owned or had access to a mobile phone; and (4) had at least two months of TB treatment remaining. Individuals with extra-pulmonary forms of TB or drug-resistant TB were eligible for inclusion. Recruitment lasted from April 13, 2018 until December 20, 2019, inclusive.

### Study Oversight and Procedures

Our trial was approved by the institutional review board of Kenyatta National Hospital and the University of Nairobi. Trial participants or their parents or guardians provided written informed consent. The trial’s protocol and statistical analysis plan were registered in advance with ClinicalTrials.gov (#NCT04119375) [17].

Our study included four treatment arms: control, SMS, platform, and Keheala. Randomization to each group was stratified by clinic, language, HIV-coinfection, and gender. Within strata, we randomized individuals into a treatment group via proportional block-randomization at a ratio of 4:3:12:12 for control, SMS, platform, and Keheala.

We hired seven study team members from Nairobi. All team members underwent training in advance of this RCT on ethical conduct of research, including confidentiality. We also trained the study team on TB disease, its treatment, barriers to medication adherence and treatment completion, and relevant behavior change strategies from the social sciences (discussed below). Members of the study team were subsequently responsible for training clinic county and sub-county coordinators on the study and enrollment procedures, and then for providing support for individuals in the Keheala intervention.

We recruited and trained clinicians on the study as follows. Prior to the study, our study team attended the National TB Program’s county and sub-county quarterly review meetings to provide information about the study and training on enrollment procedures. These meetings are typically attended by county and sub-county coordinators (supervisors). Our training took approximately 30 minutes, after which we registered supervisors willing to participate. These supervisors were provided with training materials and consent forms, and given permission to train and register their subordinates. Registration could be done via a mobile phone, by adding the provider’s clinic, name, and number.

Participants were recruited for the study by the health care providers who had been trained and registered by their supervisors. Recruitment was performed at the clinic, at the conclusion of a visit. For all individuals selected by clinicians for participation, the clinician described the study, and checked if individuals were eligible for the trial. For those individuals interested in participating in the study, clinicians obtained written consent. Then, the individual was centrally randomized into an intervention by the system.

We randomly selected a subsample of participants with drug-sensitive tuberculosis (as drug-resistant participants typically do not receive isoniazid) in the Keheala and control group, in a 3:1 ratio, for unannounced tests for the presence of isoniazid in their urine using IsoScreen by GFC Diagnostics (Oxfordshire, UK). Tests were administered by study team members who made unannounced visits to the participant’s clinic, waiting there for up to one week. If the participant was not encountered, then the team attempted to reach the participant at home the following week.

### Intervention Groups

The study included a control group that received the standard of care, plus three intervention groups, each of which received the standard of care plus a DAS intervention. Many features of the interventions were designed with behavioral science principles aimed at fostering individuals’ desire to protect not only themselves, but also others from TB [8, 18].

### Control Group

The control group received the standard of care, without further support. Individuals assigned to the control group were sent a single SMS thanking them for consenting, and informing them they would not receive any further messages.

### SMS Group

The first intervention group received a daily one-way SMS reminder that read, “Please take the medication on time.” The default time of the SMS messages was 7 a.m., as, in the standard of care, most individuals are instructed to take their TB medications in the morning [16]. The timing could be changed by the enrolling care provider and individuals could request a time change.

### Platform Group

The second intervention was a digital platform which was accessed via a protocol known as Unstructured Supplementary Service Data (USSD) and included the following features.

Each day, the platform sent an automated request that individuals self-verify medication adherence (i.e. two-way SMS); again the timing defaulted to 7 a.m., but could be changed by the enrolling care provider or the individual. The request read, “It’s time! Have you taken your medication? Dial *694# to verify.”. When individuals logged in using the short code, they were greeted by a screen requesting that they verify adherence. Individuals who failed to verify within an hour received up to two more automated reminders, at hourly intervals.

After verifying, individuals were taken to a home menu from which they could access two additional platforms features: a page with TB information, and an anonymous leaderboard-style ‘adherence contest’ in which individuals could compare their verification rate to others’ and obtain badges if they reached 90% adherence. Individuals could also access these features at any time by logging in to the platform with their short-code.

Finally, once a week, individuals received a motivational message such as, “take your medication, not so you get better but so you protect your family and friends,” or “80% of patients took their medication today, join them.”

### Keheala

The third and final intervention, Keheala, was comprised of the previously described platform along with added support by the study team, which was aimed at addressing barriers to adherence and treatment completion, and facilitating communication with individuals’ providers. Specifically, if, within an hour of sending the third request for verification, individuals still did not verify, the protocol called for subjects in the Keheala intervention group to be placed on a queue for contact by a member of the study team via either SMS or phone call. Individuals could also initiate an asynchronous conversation with the study team via a chat feature to, e.g., request a phone call, ask questions about treatment, raise concerns, or obtain assistance communicating with clinic staff.

### Deviations from Protocol

Crucially, two coding errors meant that the platform and Keheala interventions were not experienced according to protocol. The first error prevented some individuals from verifying their participation. This error affected participants in both groups from the first day of recruitment (April 13, 2018) until it was corrected on or about September 3, 2019.

The second error prevented individuals who had failed to verify from being included in the study team’s contact queue and, thus, from being contacted as frequently as required by the protocol. This error affected participants in the Keheala group from the first day of recruitment until it was corrected on or about October 24, 2019, approximately two months before the end of recruitment.

These implementation errors can be conceptualized as capacity constraints. The first constrained the number of participants who could successfully verify at a given point in time, while the second constrained the number of participants who could receive outreach on a given day. These constraints limited treatment exposure, but had no effect on assignment. See Appendix F for a detailed discussion. Separately, there was a third coding error related to treatment assignment: the string used to block-randomize participants was initially truncated. This resulted in too few patients being assigned to the Keheala group. This issue affected randomization from the first day of recruitment until it was corrected on February 20, 2019. Following this date, the share of participants randomized to Keheala increased.

### Study Outcomes

Our co-primary outcomes were: unsuccessful treatment outcome and LTFU. For the control and Keheala groups, we also considered death as a primary outcome. Death was only considered for these groups because we felt it merited a smaller minimum detectable effect (of a 1 percentage point reduction in absolute risk), and, thus an increase in the sample size. In accordance with Kenyan guidelines [15], LTFU occurs when a patient interrupts treatment for two consecutive months; death is defined as all cause mortality during the course of treatment; unsuccessful treatment outcome is a composite of LTFU, death, and treatment failure, which occurs if an individual’s sputum smear or culture is positive at month five or later. We obtained outcome data from the National TB program’s electronic database known as TIBU [15]. We obtained the outcome data on March 14, 2021, which was roughly 15 months after the final patient was recruited, and thus provided ample time for treatment completion and outcome reporting.

For individuals who participated in unannounced isoniazid urine tests, we considered medication adherence as a secondary outcome. It was defined based on the results of the IsoScreen using standard methods [19, 20].

In addition to these outcomes, for individuals in the platform and Keheala interventions, we also considered their verification rates, defined as the proportion of days on which they self-verified adherence.

### Cost Methods

We estimated the incremental cost in 2021 USD per additional participant completing treatment for each intervention compared to the control. In addition, we estimated the cost per loss to follow-up averted for each intervention compared to the control, and per death averted for Keheala compared to the control. As Keheala is a proprietary intervention, we used the cost offered to customers on a per patient-month basis as the cost of this intervention (i.e., $2.37 USD). For the costs of SMS ($0.45 USD per month) and platform ($0.80 USD per month), we estimated the relative costs of these interventions to Keheala through a detailed microcosting exercise. We considered costs only directly related to the intervention and did not include subsequent costs, such as costs of increased clinician time or medications associated with improved adherence. Further details on the cost analysis are offered in Appendix A and Appendix C.

### Statistical Analysis

Our target group sizes were 2,000 for the control group, 1,500 for the SMS group, 6,000 for the platform group, and 6,000 for the Keheala group. These targets were chosen to achieve statistical power of 90 percent with *α* = .05 for the following comparisons: 1 percentage points between the Keheala group and the control group in deaths and unsuccessful outcomes; 3.5 percentage points between the Keheala group and the SMS group in unsuccessful outcomes; and, 2.5 percentage points between the Keheala group and the platform group in unsuccessful outcomes. We did not attempt to power the study to differentiate between the SMS and platform group, or to differentiate between the SMS group and the control.

We focus on a modified intent-to-treat (mITT) population which, per our protocol, excluded the following individuals: those who were transferred out of their clinic, or whose treatment outcome could not be determined.

Consistent with our protocol, our primary analyses were as follows.

For each of our co-primary outcomes, we calculated the difference in outcomes between intervention and control group, and presented confidence intervals and p-values from two-sided, two-sample Z tests of proportions. We compared the performance of our interventions to one another in the same manner.

We also estimated the impact of our three interventions on our co-primary outcomes from two sets of least squares regressions. The first set (‘partially adjusted’) includes controls for clinic fixed effects and month of recruitment. The inclusion of month of recruitment absorbs variance in treatment outcomes over time that might otherwise have been attributed to intervention assignment, particularly due to the change in frequency of assignment following the correction of the block-randomization string.

The second (‘fully adjusted’) also controls for individual and disease characteristics. We included all individual and disease characteristics available when an individual commences treatment except height and weight, as these were often recorded with typographical errors or were missing. The characteristics included were: language preference, sex, age, HIV status, nutritional support, presence of any comorbidities, whether the patient was treated for pulmonary or extrapulmonary TB, whether the patient was being retreated for TB, whether the patient’s diagnosis was bacteriologically confirmed, and months on treatment at the time of enrollment in the study. Clinic, language preference, sex, and HIV Status were provided by the individual’s provider at the time of enrollment in the study. The remaining characteristics were recorded in TIBU or calculated from data that originated therein.

Our secondary analyses focus on individuals selected to participate in surprise isoniazid tests. We report the crude difference in the proportion of negative urine test results between the Keheala and control groups. We also report the results of least squares regressions of medication adherence on treatment assignment, controlling for clinic fixed effects and the same individual and disease characteristics listed above. In these analyses, we excluded individuals based on the same criteria as in our primary analyses, except that we did not exclude individuals with missing outcomes.

We performed a number of additional sensitivity and stratified analyses, including: restricting to individuals who were randomized within their first month of treatment, estimating the share of individuals who experienced a non-zero effect of each intervention, predicting the effect of each intervention had it been delivered to eligible individual in participating clinics – and in Kenya – during our study period, reviewing first-week verification rates before and after the first implementation error was repaired, and reviewing the relationship between the number of days on which patients were not contacted per protocol and their outcomes. These and other sensitivity analyses are described in detail in the Supplement. All analyses were performed in StataMP (versions 15 and 17), except for those in Appendix A, Appendix C, and Appendix J, which are described in detail therein.

## Results

### Patient Characteristics

We randomized 16,753 individuals, of whom 558 and 1,233 were excluded post-randomization due to misdiagnosis or transfer from their original treatment facility and missing treatment outcome data, respectively. After excluding these individuals, the mITT population comprised 14,962 individuals. Participants were similar to eligible individuals in participating clinics and in Kenya during the study period (Tbl. 1. 1,997 individuals were assigned to the control, 1,475 to the SMS group, 6,057 to the platform group, and 5,433 to the Keheala group. All groups were similar (Tbl. 2). Only 2 patients with Dr-TB were in our mITT population.

**Table 1:** Individual and Disease Characteristics for Individuals with TB During the Study Period.

|  | Kenya |  | Participating Clinics |  | This Study |  |  |  |
| --- | --- | --- | --- | --- | --- | --- | --- | --- |
|  |  |  |  |  | Randomized |  | mITT |  |
| N | 119,653 |  | 55,084 |  | 16,753 |  | 14,962 |  |
| Male (N, %) | 75,597 | 63.2 | 34,451 | 62.5 | 10,393 | 64.4 | 9,514 | 63.6 |
| Living with HIV (N, %) | 32,077 | 26.8 | 15,412 | 28.0 | 4,213 | 25.1 | 3,981 | 26.9 |
| At Least One Comorbidity (N, %) | 2,507 | 2.1 | 1,425 | 2.6 | 524 | 3.1 | 497 | 3.3 |
| Extrapulmonary (N, %) | 17,558 | 14.7 | 8,612 | 15.6 | 1,997 | 11.9 | 1,794 | 12.0 |
| Retreatment (N, %) | 9,072 | 7.6 | 4,645 | 8.4 | 1,314 | 7.8 | 1,262 | 8.7 |
| Nutrition Support (N, %) | 117,217 | 98.0 | 55,046 | 99.9 | 16,125 | 96.3 | 14,957 | 100.0 |
| Bacteriologically Confirmed (N, %) | 78,375 | 65.5 | 38,498 | 69.9 | 12,074 | 72.1 | 11,243 | 75.1 |
| Age in Years (Avg., Min–Max) | 34.6 | 0–120 | 33.9 | 0–120 | 34.4 | 0–120 | 34.4 | 0–99 |
| Misdiagnosed or Transferred Out (N, %) | 6,382 | 5.3 | 4,170 | 7.6 | 558 | 3.3 | 0 | - |
| Missing Outcome or New Case (N, %) | 47,266 | 39.5 | 6,191 | 11.2 | 1,233 | 7.4 | 0 | - |

Summaries of individual and disease characteristics for: all individuals with TB in Kenya during the study period who met inclusion criteria (“Kenya”); all individuals treated at participating clinics (“participating clinics”); all individuals who participated in the study (“this study, randomized”), and all individuals who met our preregistered inclusion criteria (“this study, mITT”). For indicator variables, we present Ns and percents. For the remaining variables, we present the mean, minimum and maximum value. Known duplicates are omitted from all columns.

**Table 2:** Individual and Disease Characteristics, by Intervention Group.

|  | Control |  | SMS |  | Platform |  | Keheala |  |
| --- | --- | --- | --- | --- | --- | --- | --- | --- |
| N | 1,997 |  | 1,475 |  | 6,057 |  | 5,433 |  |
| English (N, %) | 814 | 40.8 | 606 | 41.1 | 2,525 | 41.7 | 2,262 | 41.6 |
| Male | 1,280 | 64.1 | 936 | 63.5 | 3,857 | 63.7 | 3,441 | 63.3 |
| Living with HIV | 525 | 26.5 | 390 | 26.8 | 1,624 | 27.0 | 1,442 | 26.8 |
| At Least One Comorbidity | 75 | 3.8 | 46 | 3.1 | 202 | 3.3 | 174 | 3.2 |
| Extrapulmonary | 246 | 12.3 | 163 | 11.1 | 716 | 11.8 | 669 | 12.3 |
| Retreatment | 130 | 6.7 | 122 | 8.5 | 520 | 8.8 | 490 | 9.3 |
| Nutrition Support | 1,996 | 99.9 | 1,474 | 99.9 | 6,055 | 100.0 | 5,432 | 100.0 |
| Bacteriologically Confirmed | 1,483 | 74.3 | 1,116 | 75.7 | 4,553 | 75.2 | 4,091 | 75.3 |
| Age in Years (Avg., Min.–Max.) | 34.2 | .17–99 | 34.9 | .17–90 | 34.1 | .08–98 | 34.8 | .08–98 |
| Month of Treatment at Start | 1.2 | 0–9.57 | 1.2 | 0–9.74 | 1.2 | 0–9.97 | 1.2 | 0–9.7 |

Summaries of individual and disease characteristics by intervention group. For indicator variables, we present Ns and percents. For the remaining variables, we present the mean, minimum and maximum value.

### Primary Outcomes

Tbl 3 presents primary outcomes by intervention group. In the control group, 247 individuals (12.4%) experienced an unsuccessful outcome, 128 (6.4%) were LTFU, and 105 (5.3%) died. Relative to the control, the Keheala intervention reduced the absolute risk of unsuccessful outcomes by 2.6 percentage points (95% C.I.: 1.0-4.2; p=.001); the platform and SMS interventions both reduced the absolute risk of unsuccessful outcomes by 1.9 percentage points (SMS 95% C.I.: -0.1-4.0, p=.067; platform 95% C.I.: 0.3-3.4, p=.019). Nearly all of these reductions came from reductions in LTFU. When compared to SMS and platform, Keheala’s reductions in unsuccessful outcomes and LTFU were not significant (see Appendix B). Finally, the Keheala intervention reduced the absolute risk of death by 0.3 percentage points (95% C.I.: -0.8-1.5; p=.573). The results remain consistent when partially or fully adjusted.

### Secondary Outcomes

731 eligible participants completed surprise isoniazid urine tests for medication adherence, of whom 194 were in the control group and 537 were in the Keheala group. In the control group, 12.4% of individuals’ test results were negative. In the Keheala group, 4.8% of individuals’ results were negative, a difference relative to the control of 7.5 percentage points (95% C.I.: 2.6-12.5; p=.003).

Verification rates were 9.5% on average for individuals in the platform group and 38.2% in the Keheala group, a difference of 28.7 percentage points (95% C.I.: 27.7-29.6; p<.001).

### Cost Effectiveness

We estimated that the average cost per patient was $2.48 for SMS, $4.48 for platform, and $13.11 for Keheala. Compared to the control group, the cost per additional participant successfully completing treatment was $131 (95% C.I.: $601240) for SMS, $236 (95% C.I.: $128-2240) for platform, and $504 (95% C.I.: $305-1311) for Keheala. Additional estimates of the cost per loss to follow-up averted and death averted and estimates comparing Keheala to SMS and platform are in Appendix C.

### Sensitivity Analyses

Our results were robust to assumptions about unknown outcomes, errors in TIBU outcome data, as well as within stratified analyses on bacteriological confirmation, site of disease, and previous treatment history. See Appendix D–Appendix I

When only considering individuals who were randomized within their first month of treatment, the reduction in risk of an unsuccessful outcome was 3.8 percentage points for Keheala (95% C.I.: 1.4-6.1; p<.001), 2.8 percentage points for platform (95% C.I.: 0.5-5.2; p=.014) and 3.7 percentage points for SMS (95% C.I.: 0.7-6.7; p=.018; see Appendix G).

We estimated that 35.4% of individuals in Keheala, 15.2% of individuals in platform, and 8.7% of individuals in the SMS group experienced a non-zero individual benefit from the intervention (see Appendix J).

Had our interventions been delivered to all eligible individuals in participating clinics during our study period, we predict a reduction in risk of unsuccessful outcomes of 2.6% for Keheala, 1.9% for Platform, and 2.0% for SMS. Had our interventions been delivered to all eligible individuals in Kenya during our study period, we predict a reduction in risk of unsuccessful outcomes of 2.6% for Keheala, 1.8% for Platform, and 2.3% for SMS. See Appendix J.

Comparing first week verifications and first month contact for individuals who enrolled in the trial in the month before versus after the implementation error were repaired, we find post-repair increases of 10.3 percent and 84.2 percent, respectively. The more often the study team did not contact the patient per protocol – defined as the number of days that a patient was not contacted per protocol divided by the number of days that they should have been contacted – the worse the participants’ outcomes: 88.7% for those above the median vs. 95.2% for those below the median (diff: 6.5%; CI: 4.8-8.1%; p<0.001). See Appendix F.

## Discussion

In this trial, all three digital health interventions were associated with improvements in treatment outcomes–of 2.6 percentage points for Keheala, and 1.9 percentage points for the SMS and platform interventions–relative to an absolute risk of an unsuccessful outcome in the control of 12.4%. The bulk of these improvements came from reductions in LTFU. The interventions had a more pronounced impact among individuals initiating the intervention in the first month of treatment. Particularly for Keheala, these results are robust to statistical controls for clinic-level fixed effects as well as disease and individual characteristics, and persist in a number of sensitivity analyses with unfavorable assumptions.

Additionally, the Keheala intervention reduced medication non-adherence by 7.5 percentage points. This result is also robust to statistical controls. Although medication adherence contributes to treatment success, it is of independent interest, since higher adherence also reduces the risk of developing DR TB [21] and of disease recurrence [22].

For all three interventions in this trial, the additional costs associated with each intervention per additional participant completing treatment are likely to be more than offset by costs associated with loss to follow-up. In Kenya, the total cost to the health system and individuals per person lost to follow-up is approximately $2437 (2021 USD) [23], significantly more than costs per loss to follow-up averted for each intervention in this trial. Additionally, premature mortality due to TB is associated with multiple disability-adjusted life years [24].

Our results contribute to a nascent literature testing the impact of DAS interventions on TB treatment outcomes [21, 25, 26, 9, 27, 28, 29, 30, 31, 32, 33, 34], and may inform treatment of other diseases, such as HIV, with long-lasting treatment and for which adherence is critical not only for improving health outcomes, but also for preventing transmission and drug-resistance. These results build upon our prior RCT [9] to demonstrate the potential of the Keheala intervention to reduce unsuccessful outcomes and deaths, this time at larger scale, with a diverse patient population. They also demonstrate the potential of two additional interventions that are less resource intensive.

Our trial has a number of limitations. One is the exclusion of individuals who do not have access to or know how to operate a mobile phone, which, because these individuals are at relatively high risk of unsuccessful outcomes [35, 19, 32], may exacerbate inequality [36, 10]. A second limitation relates to our recruitment strategy, which relied upon health care providers to offer the intervention to patients: if providers chose not to participate in the study, then, necessarily, their patients were excluded and the reason for exclusion was not recorded. Participants’ similarity to eligible individuals in participating clinics and in Kenya allays this concern somewhat, as does the similarity of participants’ estimated treatment effects to predicted treatment effects for these populations. However, the study’s results should still, in our view, be seen as more representative of a program that relies on providers to recommend DAS interventions, than of a program in which patients are automatically enrolled in a DAS intervention upon diagnosis. A third limitation is that implementation errors prevented those enrolled in the platform and Keheala interventions from verifying adherence and, in the case of Keheala, from receiving as much human support as intended. Because these implementation errors operated like binding capacity constraints on verification and outreach, our estimates reflect the effects of these interventions under constrained delivery rather than under full per protocol capacity. Nonetheless, our results are consistent with existing research [3, 4, 37] showing that human support conveys benefits above-and-beyond purely automated interventions: participants in the Keheala group experienced the greatest improvements in treatment outcomes, and the repair of the implementation errors led to an increase in per protocol contacts, which are in turn associated with better treatment outcomes. Fourth, individuals were enrolled throughout their treatment, whereas, at scale, individuals would likely be enrolled at the beginning of treatment. The results for our patients enrolled early in treatment suggest that if the interventions were scaled in this way, they may, in fact, yield larger impacts than those we found in our study. Fifth, we relied on program outcome data, but extensive sensitivity analyses around this point again showed consistent and robust results. Sixth, our interventions were designed for and tested within the Kenyan context. Our results may not generalize to other settings; before being applied elsewhere, our interventions must be adapted to the local context.

The trial’s strengths include: the use of a pragmatic design in a large number of clinics, serving a diverse population of persons with TB; integration of our digital interventions with standard care without the need for specialized clinical settings; the involvement of research tools from across a diversity of fields and the use of behavioral science principles to design the interventions, specifically observability [38, 8, 18] which motivated our requests for verification, eliminating plausible deniability [8] which motivated our use of repeated reminders and follow-up by supporters for non-verifiers, and communicating a norm of adherence [8, 18] which motivated our widespread use of prosocial and normative frames.

In summary, we believe our interventions could be considered as a supplement to the standard of care, especially in resource-constrained regions where in-person support is impractical.

## Data Availability

Deidentified data produced in the present study are available upon request to the authors, and will be made available online at time of publication.

## Appendix A. Costing Estimation Methods

Keheala is a proprietary intervention, available to customers at various price-points. For our costing analysis, we used the cost per patient borne by customers such as local or national TB programs. We selected the price point offered in Kenya of 31,200,000 Kenyan Shillings or $283,920 USD (2021 exchange rate of 0.0091 USD per Kenyan Shilling) per 120,000 patient-months of treatment (or 20,000 patients completing 6 months of drug susceptible tuberculosis treatment). This is equivalent to an overall cost of $2.37 USD per patient month of treatment. We selected this price point as it is a realistic target in a country that diagnoses 80,000 cases of tuberculosis each year. Costs per patient would be higher for customers enrolling fewer patients, while lower for customers enrolling comparatively higher patients.

Since the platform and SMS interventions are not offered separately through Keheala, to estimate the incremental cost-effectiveness of each intervention arm compared to the control arm, we conducted an extensive micro-costing exercise. We used the same micro-costing approach to estimate the costs for Keheala, platform and SMS interventions used throughout the study. This decision was made as cost efficiencies realized in real world implementation won’t be realized in a clinical trial. This microcosting exercise reflected costs of patient support by these digital tools, including all materials, equipment, training, and personnel time. However, all research-related costs as well as those directly referable to standard patient care (e.g., medications, physician/nurse time) were excluded. We assumed costs associated with the standard of care (i.e., control arm) would be identical in each group. We assumed that the proportional difference in costs between the Keheala, platform, and SMS interventions estimated through micro-costing would also hold true when offered to customers outside of a clinical trial (i.e., if in the microcosting exercise, platform was 50% cheaper than Keheala, it could be offered to customers at 50% of the price of Keheala).

Throughout the study, from inception to completion of all patient follow-up, all materials, capital expenditures, and personnel time dedicated to intervention implementation and delivery were carefully recorded. For materials and capital expenditures, we attributed costs to each intervention group based on level of effort (i.e., number of participants in each arm and use of these items for implementation and delivery of the intervention). Materials and capital expenditures included items such as technology development, mobile airtime, office broadband, technical support, and patient information cards. For personnel, every day all team members self-reported their working hours and attributed them to one of five categories: Keheala-specific support, research, meetings, secondary roles, and other. At the end of the study, these hours were tallied and multiplied by staff-specific salaries to arrive at an overall cost. All personnel time associated with Keheala-specific support was attributed to the Keheala arm, while personnel time associated with meetings, secondary roles, and other tasks were proportionally allocated between Keheala, platform, and SMS arms based on the number of participants randomized to each arm (Tbl. S.I.1).

## Appendix B. Comparison of Keheala to Platform and SMS

In Tbl S.I.5, we present the difference in the reduction of risk of unsuccessful outcomes and LTFU, between Keheala and the platform intervention, and betwenn Kehealla and the SMS interventions. Confidence intervals and p-values are from unadjusted tests of proportions.

## Appendix C. Costing Analysis Methods

Tbl. S.I.2 displays the information used to perform the cost-effectiveness analysis, where we estimate the incremental cost-effectiveness of each intervention compared to standard of care for the outcome of successful treatment completion. We calculated the number of patient months on treatment for all participants in the trial and calculated the average number of patients months on treatment per patient to arrive at an average cost of each intervention per patient. We then used calculated differences in the proportion of people completing treatment between each intervention arm and the control (see Tbl. 3 in the main text) to calculate the incremental cost per additional person completing treatment.

**Table 3:** Unsuccessful Outcomes, LTFU, and Deaths by Intervention.

|  | Absolute Risk |  | Reduction in Risk Relative to the Control |  |  |  |  |  |  |  |  |
| --- | --- | --- | --- | --- | --- | --- | --- | --- | --- | --- | --- |
|  | N | % | Unadjusted |  |  | Partially Adjusted |  |  | Fully Adjusted |  |  |
|  |  |  | % | 95% C.I. | p-value | % | 95% C.I. | p-value | % | 95% C.I. | p-value |
| **Control (N=1,997)** |  |  |  |  |  |  |  |  |  |  |  |
| Unsuccessful Outcomes | 247.0 | 12.4 |  |  |  |  |  |  |  |  |  |
| LTFU | 128.0 | 6.4 |  |  |  |  |  |  |  |  |  |
| Death | 105.0 | 5.3 |  |  |  |  |  |  |  |  |  |
| **SMS (N=1,475)** |  |  |  |  |  |  |  |  |  |  |  |
| Unsuccessful Outcomes | 154.0 | 10.4 | 1.9 | (-0.1–4.0) | .067 | 1.5 | (-0.6–3.6) | .158 | 1.8 | (-0.3–4.0) | .097 |
| LTFU | 70.0 | 4.7 | 1.7 | (0.3–3.1) | .020 | 1.6 | (0.2–3.0) | .027 | 1.6 | (0.1–3.1) | .036 |
| Death | 69.0 | 4.7 | 0.6 | (-0.9–2.1) | .443 | 0.1 | (-1.4–1.6) | .873 | 0.3 | (-1.2–1.9) | .657 |
| **Platform (N=6,057)** |  |  |  |  |  |  |  |  |  |  |  |
| Unsuccessful Outcomes | 637.0 | 10.5 | 1.9 | (0.3–3.4) | .019 | 1.8 | (0.2–3.3) | .027 | 2.2 | (0.5–3.8) | .009 |
| LTFU | 271.0 | 4.5 | 1.9 | (0.9–3.0) | <.001 | 2.0 | (0.9–3.0) | <.001 | 2.2 | (1.1–3.3) | <.001 |
| Death | 323.0 | 5.3 | -0.1 | (-1.2–1.0) | .895 | -0.3 | (-1.4–0.9) | .641 | -0.2 | (-1.3–1.0) | .760 |
| **Keheala (N=5,433)** |  |  |  |  |  |  |  |  |  |  |  |
| Unsuccessful Outcomes | 530.0 | 9.8 | 2.6 | (1.0–4.2) | .001 | 2.7 | (1.1–4.3) | <.001 | 3.2 | (1.5–4.8) | <.001 |
| LTFU | 211.0 | 3.9 | 2.5 | (1.5–3.6) | <.001 | 2.6 | (1.5–3.6) | <.001 | 2.7 | (1.6–3.9) | <.001 |
| Death | 268.0 | 4.9 | 0.3 | (-0.8–1.5) | .573 | 0.2 | (-0.9–1.4) | .672 | 0.5 | (-0.7–1.6) | .451 |

Absolute risk of each primary outcome in each group, and reduction in risk from each intervention relative to the control. The modified Intent-to-treat (mITT) population included all eligible individuals who underwent randomization and were not subject to the following exclusions: transferred out, indeterminate treatment outcome. Unadjusted 95% confidence intervals and p-values are from two-sided, two-sample Z tests of proportions. Adjusted coefficients are from linear regressions of the primary outcome on intervention assignment. Partially adjusted models include clinic and month-year fixed effects. Fully adjusted models add controls for the following individual characteristics: language preference, sex, age, HIV status, nutritional support, presence of any comorbidities, whether the patient was infected with pulmonary or extrapulmonary TB, whether the patient was being retreated for TB, whether the patient's diagnosis was bacteriologically confirmed, and months on treatment at the time of enrollment in the study. Unadjusted and partially adjusted analyses include all individuals. Fully adjusted models include 13,617 individuals for whom individual characteristics were available.

**Table 4:** Medication Non-adherence in the Control and Keheala Groups.

|  | Absolute Risk |  | Reduction in Risk Relative to the Control |  |  |  |  |  |  |  |  |
| --- | --- | --- | --- | --- | --- | --- | --- | --- | --- | --- | --- |
|  | N | % | Unadjusted |  |  | Partially Adjusted |  |  | Fully Adjusted |  |  |
|  |  |  | % | 95% C.I. | p-value | % | 95% C.I. | p-value | % | 95% C.I. | p-value |
| **Control (N=194)** | 24 | 12.4 |  |  |  |  |  |  |  |  |  |
| **Keheala (N=537)** | 26 | 4.8 | 7.5 | (2.6–12.5) | .003 | 6.1 | (1.0–11.2) | .020 | 6.5 | (1.5–11.6) | .012 |

Absolute risk of medication non-adherence and reduction in risk from Keheala relative to the control. The mITT population included all eligible individuals who participated in surprise isoniazid urine tests and did not transfer out. Unadjusted 95% confidence intervals and p-values are from two-sided, two-sample Z tests of proportions. Adjusted coefficients are from linear regressions of the primary outcome on intervention assignment. Partially adjusted models include clinic fixed effects. Fully adjusted models add controls for the following individual characteristics: language preference, sex, age, HIV status, nutritional support, presence of any comorbidities, whether the patient was infected with pulmonary or extrapulmonary TB, whether the patient was being retreated for TB, whether the patient's diagnosis was bacteriologically confirmed, and months on treatment at the time of enrollment in the study. Unadjusted and partially adjusted analyses include all individuals. Fully adjusted models include 687 patients for whom individual characteristics were available.

In Tbl. S.I.3, we calculate the incremental cost-effectiveness ratio of the Keheala group to the SMS and Platform groups.

In Tbl. S.I.4, we calculate the incremental cost-effectiveness ratio of the Keheala group to the standard of care for the outcome of death.

## Appendix D. Analysis of Individuals Who Transferred Out, or Missing Outcome Data

Consistent with our pre-registered protocol [17], the analyses presented in the manuscript are performed after post-randomization exclusion of 558 and 668 individuals who transferred from their original treatment facility and were missing treatment outcome data, respectively.

Tbl. S.I.6 summarizes the percent of individuals who experienced these three outcomes by intervention. As can be seen from the table, these percentages relatively equal across intervention groups.

Tbl. S.I.7 repeats our primary analysis, which appears in Tbl. 3 in the manuscript, but under the presumption that all individuals whose outcome data was missing experienced an unsuccessful outcome. The estimated impact of Keheala is very similar to that reported in the manuscript, and it remains highly statistically significant. The results for the platform and SMS interventions are weaker than those reported in our primary analyses, and are not statistically significant, or only marginally so.

Tbl. **??** repeats this exercise, but under the presumption that, in addition to those with missing outcome data, all individuals who transferred out also experienced an unsuccessful outcome. The effect of the Keheala intervention is still large and highly significant. The effect of the platform intervention is smaller than the effect reported in the manuscript, and not statistically significant, or only marginally so. The effect of the SMS intervention is further muted, and not statistically significant.

## Appendix E. Analyses of Bacteriologically Confirmed, Non-Extrapulmonary, and Non-Retreatment Individuals

We repeat our main analyses (the results of which appear in the manuscript’s Tbl. 3), but restricting to bacteriologically confirmed individuals (Tbl. S.I.8), those with pulmonary TB (Tbl. S.I.9), and those who were not being retreated (Tbl. S.I.10). For Keheala, the estimated effect is consistent with that presented in the manuscript, with the exception of unsuccessful outcomes for bacteriologically confirmed individuals; for that case, the estimated effect of Keheala is muted. For platform, the estimated effect is muted in some analyses. For SMS, the estimated effect remains consistent with the effect presented in the manuscript, but the reduced sample means it is often not statistically significant.

## Appendix F. Analysis of the Implementation Errors’ Effects on Verification and Contact

There were two implementation errors which prevented patients from experiencing their intervention according to the protocol. These errors can be understood as system-level capacity constraints that varied over time, but were orthogonal to treatment assignment and individual patient characteristics. In this appendix, we assess how verification and contact changed once the implementation errors were corrected, and capacity was thereby increased.

### Verification Implementation Error

The first implementation error prevented some patients who tried to verify from successfully doing so. The problem was described to us by the developer as one of ‘congestion’ when processing verifications: if many people attempted to verify at once, those who waited too long for their turn to be processed ‘timed out’ and were never processed. One way to conceptualize this implementation error is as a capacity constraint that restricted the number of participants who could verify at a given moment. This implementation error affected patients in the Keheala and Platform groups, beginning on the first day of recruitment (April 13, 2018) until it was repaired on or about Sept 4, 2019.

### Keheala Group

By various metrics, verification in the Keheala group increased substantially after this date. When comparing the month before the implementation error was repaired to the month after, mean verification rates jumped from 27.3 to 30.5% (diff=.032, 95% CI: 0.025-0.038, p<0.001 in a proportions test; see Fig.F.1). This difference is statistically significant for a window of any size between 10 and 150 days. When comparing all patients who registered before vs. after all who registered after September 4, 2019, we find a decrease in mean verifications rates from 38.3 to 37.1 (diff=0.012, 95% CI: 0.007-0.017, p<0.001), finding a particularly stark increase in verification rates in the first few days after a patient registered for the Keheala intervention (see Fig. F.2): verification rates on the first 2, 3, and 4 day increased by 0.046 (95% CI: 0.023-0.074, p<0.001), 0.047 (95% CI: 0.025-0.071, p<0.001), and 0.038 (95% CI: 0.018-0.059, p<0.001), respectively.

### Platform Group

Verification rates in the platform group likewise increased after the first implementation error was repaired on September 4, 2019. When comparing the month before the implementation error was repaired to the month after, mean verification rates jumped from 9.79 to 10.58% (diff=.008, 95%CI: 0.004-0.012, p<0.001 in proportion test; see Fig. F.3). This difference is statistically significant for a window of any size between 10 and 70 days. When comparing all patients who registered before vs. after all who registered after September 4, 2019, we find an increase in mean verifications rates from 12.0 to 12.5% (diff= 0.005, 95%CI: 0.002-0.008, p=0.0026). When comparing these increases to those observed in the Keheala group, it may be helpful to note that mean verification rates in the platform group are roughly a third of those in the Keheala group throughout the study period.

**Figure F.1:**
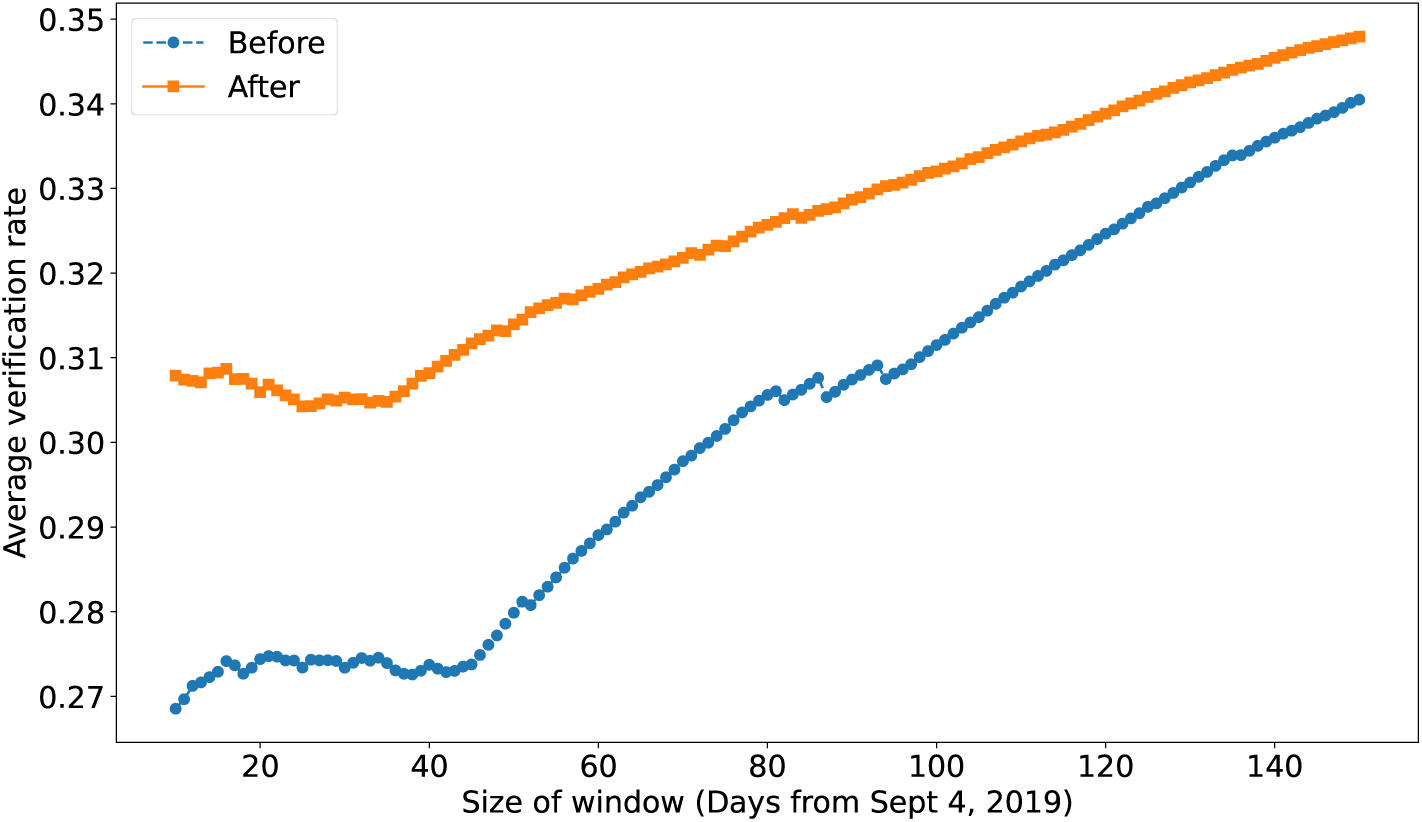
Keheala group verification rate for various time periods immediately before and after the repair of the implementation error.

### Contact Implementation Error

The second implementation error prevented some patients who should have been contacted (because they had failed to verify) from appearing on the ‘contact queue’ which the study team used to determine who to text and call on a given day – and, thus from being contacted when they should have been. From the study team’s perspective, it would typically appear that they had contacted everyone that should have been contacted on a given day, when in fact they had not been provided the complete list. This implementation error was a result of incorrect logic in the process that displayed the contact queue based on the underlying data. Unfortunately, the process did not store the list of names displayed. This implementation error can likewise be conceptualized as a capacity constraint, this time on the number of patients who could be contacted on a given day. The second implementation error affected patients from the first day of recruitment, until it was repaired on or about October 29, 2019.

By various metrics, the share of patients contacted as required by the protocol, i.e., when they had not verified in the previous 24 hours, increased substantially after this date. When comparing the month before the implementation error was repaired to the month after, the share contacted jumped from 7.0% to 17.8% (diff=0.108, 95% CI: 0.103-0.113, p<0.001 in a proportions test; see Fig. F.4). The average contact rate for all days before the implementation error was 9.6%, while it was 18.4% for all days after the implementation error was fixed (diff=0.087, 95% CI:0.084-0.090, p<0.001). Similarly, the average contact rate for those who registered before the implementation error was 10.5% and 22.8% for those who registered after (diff=0.123, 95% CI:0.111-0.134, p<0.001). When focusing on the first days after a patient registered for the Keheala intervention, we find a particularly stark increase in contact rates, ranging from 9.9% (day 2, 95% CI: 0.044-0.170, p=0.0015) to 19.2% (day 3, 95% CI: 0.133-0.257, p<0.001; see Fig. F.5).

**Figure F.2:**
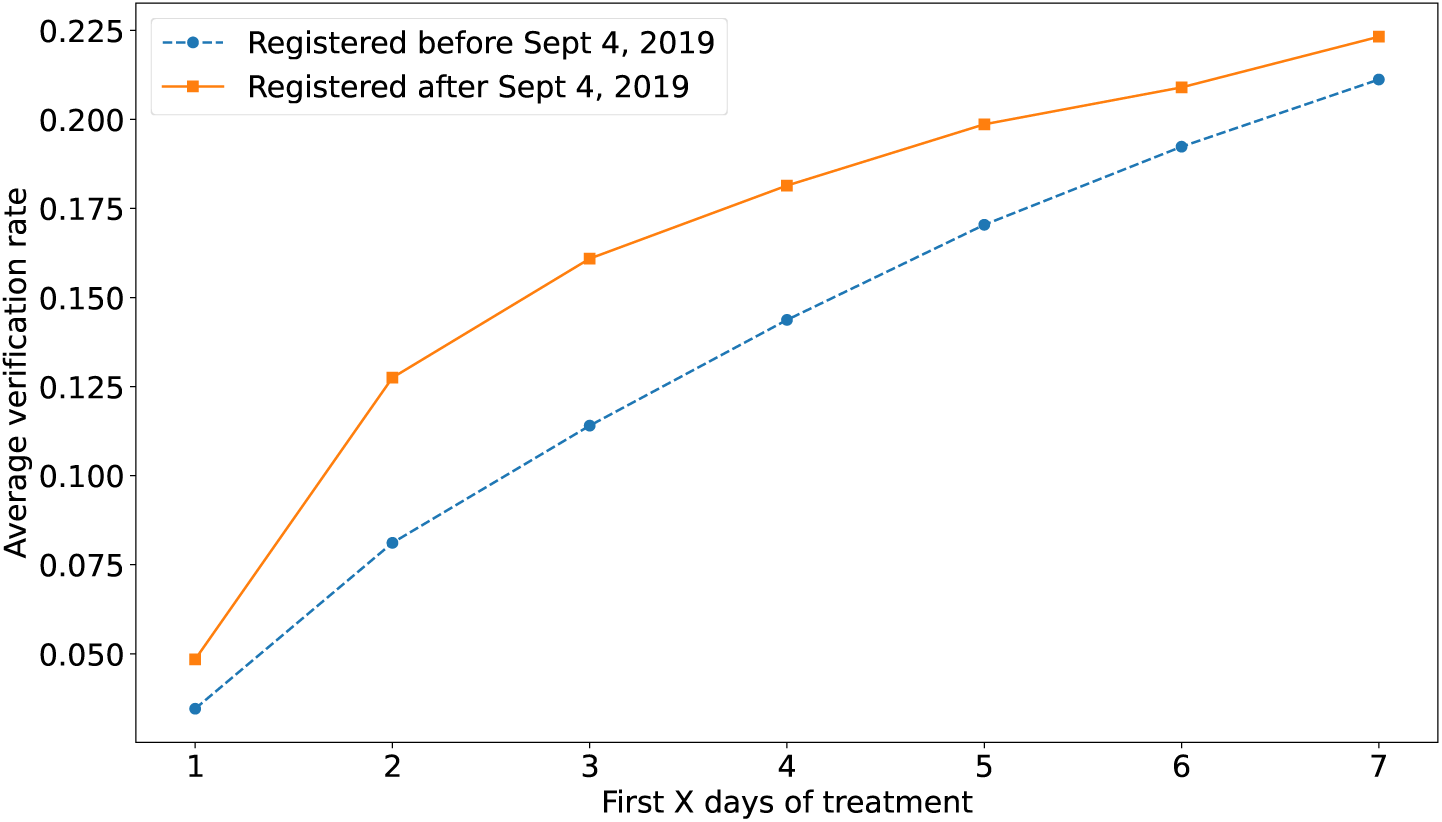
Keheala group verification rate for those that registered before and after the repair of the implementation error.

We have performed a number of analyses to try to understand whether patients whose experience of the Keheala intervention closer to protocol experienced better outcomes.

We begin by noting that we lack power to perform analyses which compare treatment outcomes of patients who enrolled in the period after the second implementation error was repaired (and who, thus, could have experienced the intervention according to the protocol; n=77) to patients in other interventions, or to patients who enrolled before this period.

Instead, we explore whether increased verification and contact were associated with improved treatment outcomes over the course of the entire study period.

**Figure F.3:**
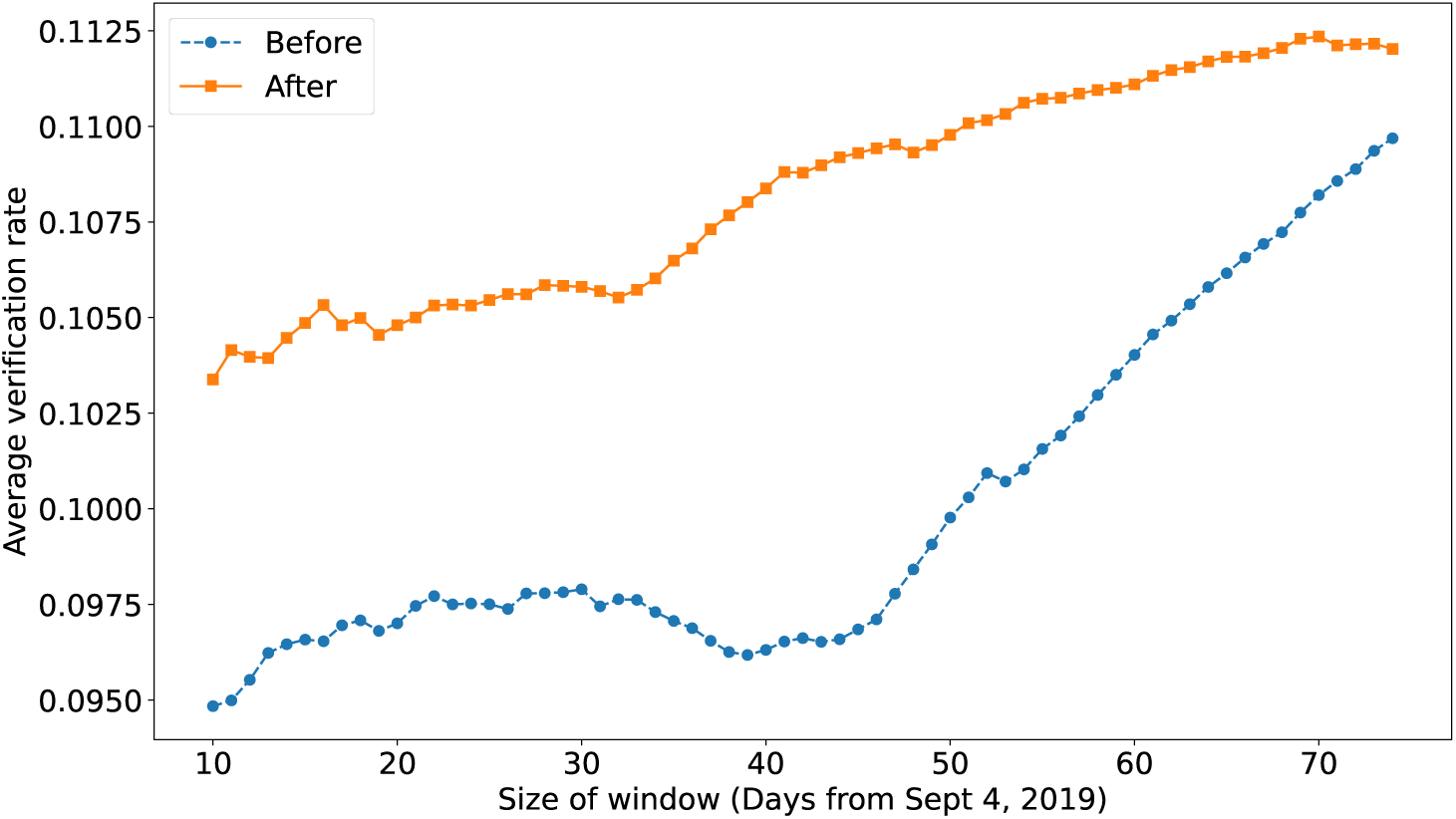
Platform group verification rate for various time periods immediately before and after the implementation error was repaired.

We find a positive association between verification and treatment outcomes. The median verification rate on a per patient basis was 30.6%. For those with a verification rate above the median, the proportion of successful outcomes was 95.9% whereas for those with a low verification rate, the proportion of successful outcomes was 88.1% (diff: 0.078, 95% CI: 0.062-0.095, p<0.001) However, we emphasize that this correlation reflects underlying differences between patients who verify more often, and should not be interpreted as representing the causal effect of verification on treatment outcomes.

One way to address this shortcoming is to focus on the period before the contact implementation error was repaired, and exploit pseudo-random variation in contact rates to see if those patients who were – to some extent due to chance – contacted more often also had higher verification rates and better treatment outcomes.

We begin by considering the effect of contact on verification. Focusing on the period before the second implementation error was repaired, and there was thus pseudo-random variation in rate at which patients were contacted when they should have been, we find that increased contact led to higher verification rates. We define the ‘missed contact rate’ as the share of days that the individual was not contacted when they should have been, according to the protocol, i.e. they were not contacted on a given day after failing to verify on the previous day. Those with a high missed contact rate (above median) had an average verification rate of 6.7 while those with a low missed contact rate had an average verification rate of 68.9 (diff=0.622, 95% CI: 0.620-0.624, p<0.001).

**Figure F.4:**
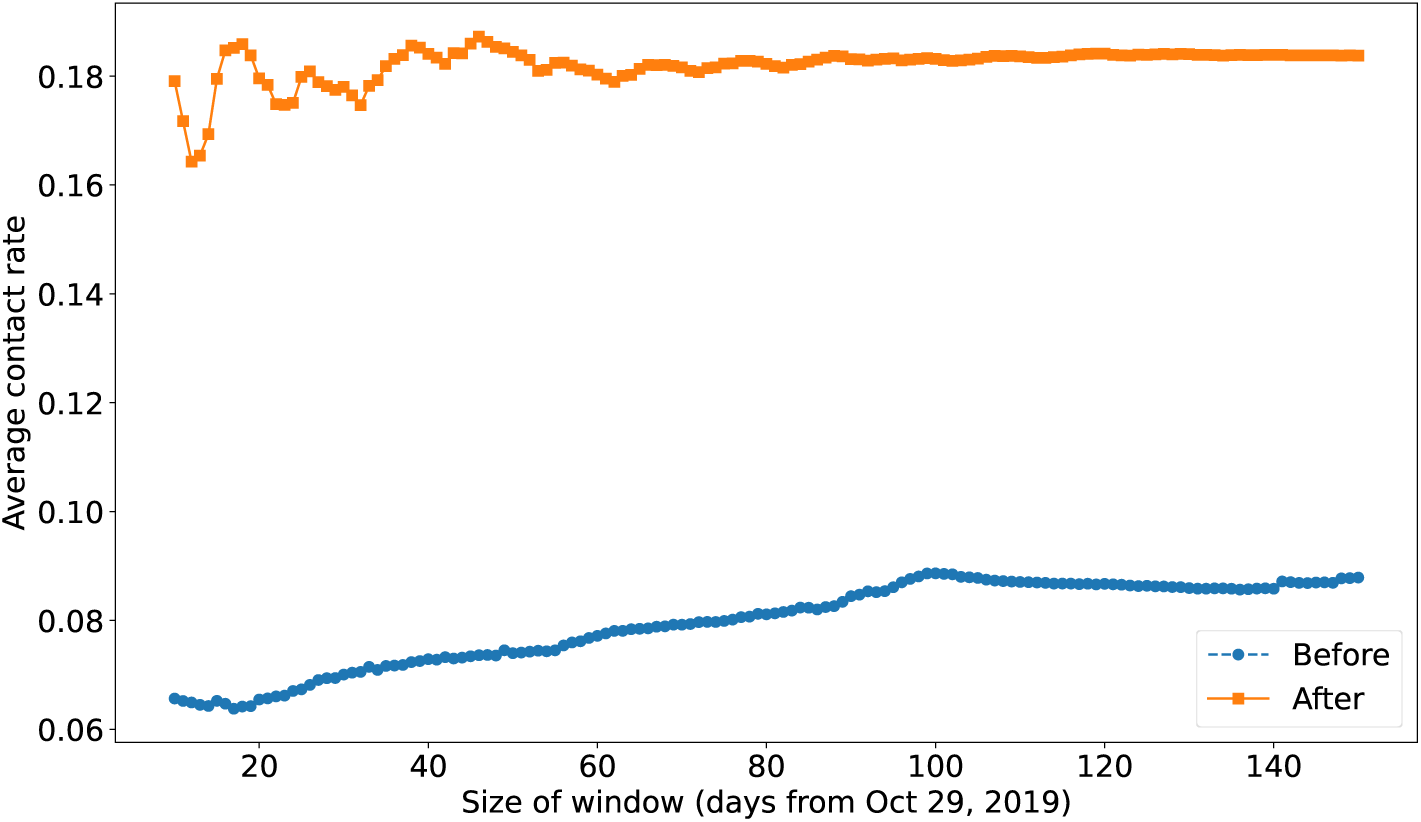
Keheala group contact rate for various time periods immediately before and after the repair of the implementation error.

In the manuscript, we reported, “We estimate that, in the absence of the implementation errors, first week verifications and first month contact would have increased 10.3 percent and 84.2percent, respectively.” These estimates are from tests of proportions comparing first week verifications and first month contacts for participants who registered for Keheala in the month preceding the implementation error’s repair on October 29, 2019 vs. for participants who registered after this date.

Next, we consider the relationship between missed contact (i.e., individuals were not contacted when they should have been) and treatment outcomes. For those with a low missed contact rate (below median), the proportion of successful outcomes was 95.2% whereas for those with a high missed contact rate, the proportion of successful outcomes was 88.7% (diff: 0.065, 95% CI: 0.048-0.081, p<0.001).

**Figure F.5:**
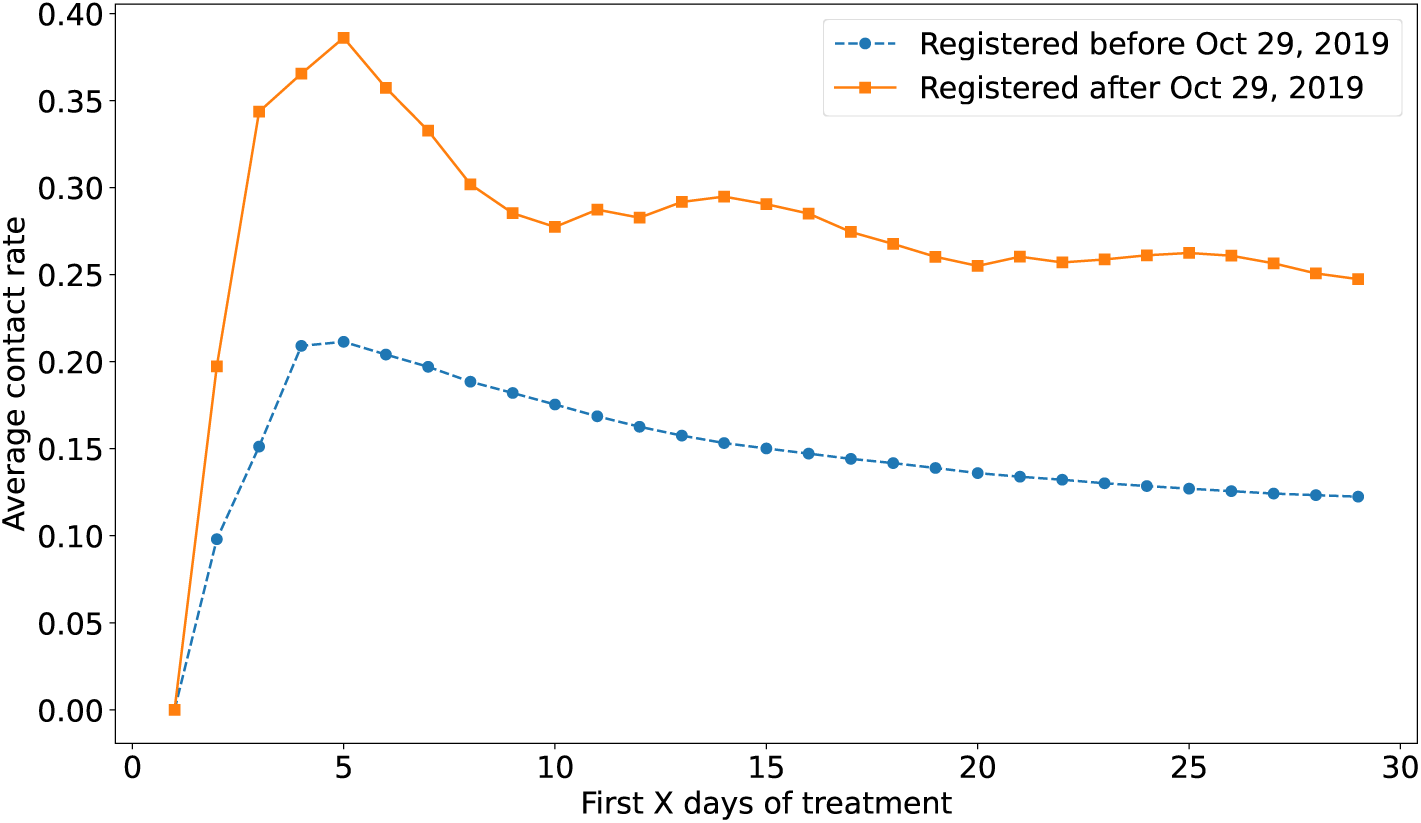
Keheala group contact rate for various time periods immediately before and after the repair of the implementation error.

## Appendix G. Sensitivity to Month of Treatment at Time of Enrollment

Participants were eligible to participate in our study so long as they had at least two months of TB treatment remaining. If, however, our interventions were implemented as part of the standard of care, most individuals would likely be enrolled in the intervention at or near the time at which their TB treatment commenced. Therefore, we consider the extent to which our interventions’ effects varied by the amount of time that had passed since they’d commenced treatment on the day on which they were enrolled. Specifically, we split the sample into those who received the intervention in their first month of TB treatment, and those who received it thereafter. Then, we repeat our analyses of our primary outcomes.

Amongst participants who commenced their intervention in the first month of treatment, unsuccessful outcomes were 15.2% in the control group, 11.5% in the Keheala group, 12.4% in the platform group, and 12.3% in the SMS group. These amount to reductions of 3.8 percentage points for Keheala (1.4–6.1; p<.001), 2.8 percentage points for platform (0.5–5.2; p=.014), and 3.7 percentage points for SMS (0.7–6.7; p=.018).

Amongst participants who commenced their intervention after the first month of treatment, unsuccessful outcomes were 16.6% in the control group, 15.3 in the Keheala group, 17.1% in the platform group, and 17.5% in the SMS group. These amount to a reduction of 1.3 percentage points for Keheala (-1.5–4.0; p=.181), and increases of 0.6 percentage points for platform (-2.2–3.3; p=.688) and 1.0 percentage points for SMS (-2.7–4.6; p=.608).

Amongst participants who commenced their intervention in the first month of treatment, LTFU was 7.5% in the control group, 4.4% in the Keheala group, 5.0% in the platform group, and 5.4% in the SMS group. These amount to reductions of 3.2 percentage points for Keheala (1.5–4.9; p<.001), 2.6 percentage points for platform (0.9–4.3; p=.001), and 2.2 for SMS (0.1–4.4; p=.050).

Amongst participants who commenced their intervention after the first month of treatment, LTFU were 4.8% in the control group, 3.2% in the Keheala group, 3.8% in the platform group, and 3.9% in the SMS group. These amount to reductions of 1.5 percentage points for Keheala (0.0–3.1; p=.020), 1.0 percentage points for platform (-0.6–2.6; p=.201), and 0.9 percentage points for SMS (-1.1– 2.9; p=.408).

Amongst participants who commenced their intervention in the first month of treatment, deaths were 6.7% in the control group, 6.0% in the Keheala group, 6.5% in the platform group, and 4.8% in the SMS group. These amount to a reduction of 0.7 percentage points for Keheala (-1.0–2.4; p=.211), 0.2 percentage points for platform (-1.5–1.8; p=849), and 1.8 percentage points for SMS (-0.2– 3.9; p=.088).

Amongst participants who commenced their intervention after the first month of treatment, deaths were 3.4% in the control group, 3.6% in the Keheala group, 3.8% in the platform group, and 4.5% in the SMS group. These amount to an increase of 0.3 percentage points for Keheala (-1.7–1.1; p=.643), an increase of 0.5 percentage points for platform (-1.0–1.9; p=.545), and an increase of 1.1 percentage points for SMS (-0.9–3.1; p=.260).

We also regress unsuccessful outcomes and LTFU on intervention assignment, separately for each of these two subsamples (see Tbls. S.I.11–S.I.12. These analyses largely corroborate the findings above: the effects of the interventions on unsuccessful outcomes and LTFU are roughly twice as strong amongst those who commence their intervention during the first month of treatment.

## Appendix H. Sensitivity to Errors in Digitization of TIBU Outcome Data

In this section, we explore the extent to which our findings are robust to errors in the outcome data. We do so by considering the impact on our analysis of various rates of type I (outcomes that are incorrectly recorded as unsuccessful) and type II errors (outcomes that are incorrectly recorded as successful).

For a given rate of type I and type II error, we adjust the rate of unsuccessful outcomes in an intervention group according to the formula: 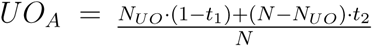 where *UO_A_* represents the proportion of unsuccessful outcomes after adjusting for type I and type II errors, *N_UO_*represents the number of unsuccessful outcomes in that group, *t*_1_ represents the type I error rate, *t*_2_ represents the type II error rate, and *N* represents the total number of individuals in the group. We then calculate the difference in unsuccessful outcomes between treatments, and the significance levels from a two-sided, two-sample test of proportions between the treatments.

In Tables S.I.13–S.I.15, we present the results for different rates of type I and type II errors.

## Appendix I. Outcome Data Quality Assurance

To test the quality of outcome data in the TIBU digital registry, we visited a subset of clinics, and obtained outcome data for participants in our study from the clinics’ paper registries–the original records that are digitized and entered into TIBU. Clinics were selected via the following cluster-randomization procedure. Within each county, we ordered sub-counties by the number of participants, and split these sub-counties into two groups at the point where 50% or more of the county’s participants were represented, so that one ‘block’ of sub-counties included a relatively small group of the largest sub-counties, and the other block included a larger group of smaller sub-counties. We then selected one sub-county from each block to represent the county, and included all clinics within that sub-county. In the end, we were able to provide outcome data from the paper records for 2,236 individuals. (Note that we excluded Wajir from the exercise because it is logistically difficult to reach and had only 34 participants.)

In Tbl. S.I.16, we present a comparison of treatment outcomes in the TIBU register with those in the paper records for these individuals. As can be seen, there are a small, but meaningful number of discrepancies; in all, an average of 2.5% of successful outcomes are mislabeled (these are type II errors, or false negatives), and an average of 7.7% of unsuccessful outcomes are mislabeled (these are type I errors, or false positives).

In Tbl. S.I.17, we present the proportion of outcomes that were mislabeled as successful or unsuccessful, broken down by treatment and the type of outcome. From this table, it can be seen that an average of only 2.5% of successful outcomes are mislabeled, whereas an average of 7.7% of unsuccessful outcomes are mislabeled. The proportion of false unsuccessful outcomes is highest for the Keheala group and the platform group: these error rates are 5.7% (p=.23) and 5.0% (p=.26) larger than the control group, respectively.

As can be seen from a comparison of the error rates in this table to those in Appendix H, most of our results are robust to error rates in excess of those we found in practice.

## Appendix J. Estimated Individual Treatment Effects of the Interventions on Primary Outcomes

In this section, we focus on the distribution of individual treatment effects on unsuccessful outcomes within each intervention (as opposed to focusing on just the average effect). Towards this aim, we trained a causal random forest model [39] which provided estimated treatment effects conditional on individuals’ personal and disease characteristics, with accompanying confidence intervals. These are estimates of the probability of an unsuccessful outcome with and without each intervention, for each individual. We included the same individual and disease characteristics in our causal forest model that we employed in our primary analyses.

We then used these estimated individual treatment effects in three ways. First, we average the estimates to obtain an alternate estimate of the average treatment effect of our interventions. This is intended as another robustness check. Second, we evaluated the heterogeneity of estimated treatment effects. Specifically, we calculated the range of estimated individual treatment effects. Third, we calculated the proportion of participants who, based on their individual estimated treatment effect and confidence interval, experienced a benefit of each intervention that was statistically greater than zero. These analyses were performed using the python libraries matplotlib, pandas, numpy, and scipy, as well as the R library grf [40].

We begin by reporting the average of the individual treatments effects. For the Keheala intervention it is 2.6 percentage points. For the platform intervention it is 1.9 percentage points, and for the SMS intervention it is 1.9 percentage points. These averages are similar to the estimated average treatment effects reported in Sec. a.

Next, we report the proportion of individuals for whom we can reject that the intervention had no effect–that is, the proportion whose estimated treatment effect is statistically distinguishable from zero because their confidence interval does not overlap with zero. In the Keheala group, 35.5% of individuals’ confidence intervals do not overlap with zero. In the platform group, 15.2% of individuals’ confidence intervals do not overlap with zero, and in the SMS group, 8.7% of confidence intervals do not overlap with zero.

Finally, we report the variation across individuals in estimated treatment effects. In the Keheala group, the estimated range is -0.6%-6.7% percentage points. In the platform group, it is -1.8%-6.5% percentage points. In the SMS group, it is -3.2%-7.8% percentage points, which is, relatively speaking, large.

Using the causal forest model, we can provide the same statistics for additional populations of interest. Had the intervention been delivered to all patients in participating clinics during the study period, we predict that the individual treatment effects would have averaged 2.6 for Keheala (range: -0.3-6.4%; 39.3% of individuals’ C.I.’s overlapping zero), 1.9% for Platform (range: -1.9-6.1%; 18.3% of individuals’ C.I.’s overlapping zero), and 2.0% for SMS (range: -2.5-6.9%; 12.6% of individuals’ C.I.’s overlapping zero). Had the intervention been delivered to all patients in Kenya during the study period, we predict that the individual treatment effects would have averaged 2.6 for Keheala (range: -0.4-6.4%; 38.0% of individuals’ C.I.’s overlapping zero), 1.8% for Platform (range: -1.9-6.1%; 19.7% of individuals’ C.I.’s overlapping zero), and 2.3% for SMS (range: -2.5-6.9%; 15.9% of individuals’ C.I.’s overlapping zero)

**Table S.I.1:** Personnel time (meetings, secondary roles, and other tasks) allocated to each treatment arm (%)

|  |  |
| --- | --- |
| Keheala | 42 |
| Platform | 47 |
| SMS | 11 |

**Table S.I.2:** Cost by Intervention.

|  | Participants | Avg. Patient Months | Cost per Month (USD) | Cost per Participant (USD) | Incremental Cost vs. SoC (per 100 patients enrolled) | Incremental Participants Completing Treatment (per 100 patients enrolled) | 95% CI | Cost-Effectiveness Ratio | 95% CI |
| --- | --- | --- | --- | --- | --- | --- | --- | --- | --- |
| Standard of Care | 1977 | — | — | — | — | — | — | — | — |
| SMS | 1475 | 5.52 | 0.45 | 2.48 | \$248 | 1.9 | (0.2–4.1) | \$131 | (\$60–\$1240) |
| Platform | 6057 | 5.60 | 0.80 | 4.48 | \$448 | 1.9 | (0.2–3.5) | \$236 | (\$128–\$2240) |
| Keheala | 5433 | 5.53 | 2.37 | 13.11 | \$1311 | 2.6 | (1.0–4.3) | \$504 | (\$305–\$1311) |

**Table S.I.3:** Incremental Cost of Keheala Relative to the SMS and Platform Interventions.

| Comparison | Incremental Cost of Keheala (2021 USD; per 100 patients enrolled) | Incremental Participants Completing Treatment (per 100 patients enrolled) | 95% CI | Cost-Effectiveness Ratio | 95% CI |
| --- | --- | --- | --- | --- | --- |
| Keheala vs. SMS | \$1063 | 0.7 | (–1.1 to 2.4) | \$1519 | (\$443 to dominated) |
| Keheala vs. Platform | \$863 | 0.8 | (–0.3 to 1.9) | \$1079 | (\$454 to dominated) |

**Table S.I.4:** Incremental Cost of Keheala Relative to the Standard of Care.

|  | Incremental Cost of Keheala (2021 USD; per 100 patients enrolled) | Deaths Averted (per 100 patients enrolled) | 95% CI | Cost-Effectiveness Ratio | 95% CI |
| --- | --- | --- | --- | --- | --- |
| Keheala vs. Standard of Care | \$1311 | 0.3 | (–0.1 to 1.5) | \$4370 | (\$874 to dominated) |

**Table S.I.5:**
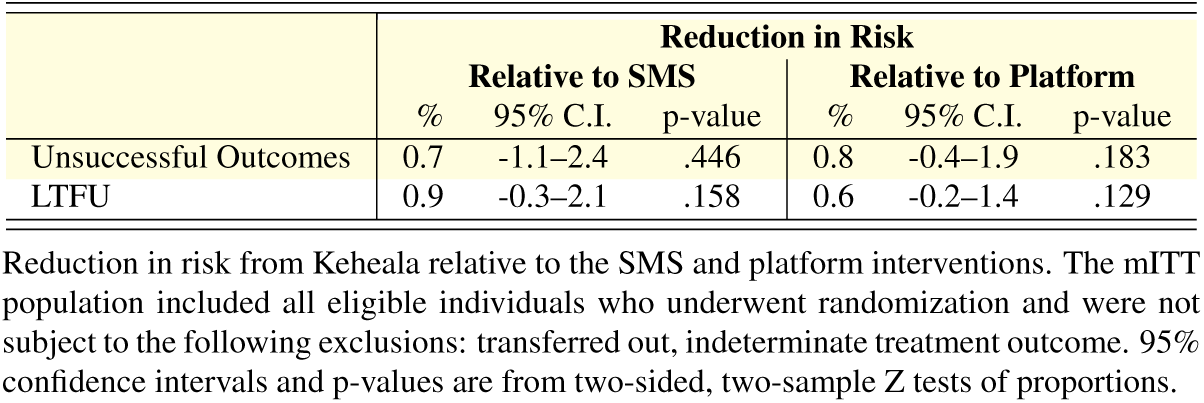
Comparison of Keheala to the SMS and Platform Interventions.

**Table S.I.6:** Misdiagnosed, Transferred Out, and Missing Outcomes by Intervention.

|  | MD | TO | N/A or Blank | NC |
| --- | --- | --- | --- | --- |
| Keheala | 0.4 | 3.5 | 3.7 | 3.3 |
| Platform | 0.3 | 3.2 | 4.2 | 3.4 |
| SMS | 0.3 | 3.7 | 4.5 | 3.5 |
| Control | 0.4 | 3.0 | 3.7 | 3.4 |
| Total | 0.4 | 3.3 | 4.0 | 3.4 |

The percentage of participants who were misdiagnosed (MD), transferred out (TO), were missing outcome data (N/A), or were marked as not completed (NC) in each of our intervention groups.

**Table S.I.7:** Regressions of Unsuccessful Outcome and LTFU on Intervention Assignment, with Missing Outcomes and MD/TO Coded as Unsuccessful Outcomes.

|  | MD/TO dropped |  |  | MD/TO coded as unsuccessful |  |  |
| --- | --- | --- | --- | --- | --- | --- |
|  | (1) | (2) | (3) | (4) | (5) | (6) |
| Keheala | -2.6**<br>(0.9) | -3.0**<br>(1.0) | -3.1**<br>(1.0) | -2.2*<br>(1.0) | -2.5*<br>(1.0) | -2.6*<br>(1.0) |
| Platform | -1.8*<br>(0.9) | -1.9*<br>(0.9) | -2.0*<br>(0.9) | -1.6<br>(0.9) | -1.6<br>(0.9) | -1.7<br>(0.9) |
| SMS | -1.4<br>(1.3) | -1.4<br>(1.4) | -1.4<br>(1.4) | -0.7<br>(1.4) | -0.5<br>(1.5) | -0.7<br>(1.5) |
| Clinic & Month Dummies |  | Yes | Yes |  | Yes | Yes |
| Individual Chars. |  |  | Yes |  |  | Yes |
| N | 15576 | 14595 | 14025 | 16134 | 15130 | 14545 |

Estimated reduction of risk (relative to the control) from linear regressions of unsuccessful outcomes on treatment assignment, assuming that individuals whose outcome data was missing experienced unsuccessful outcomes (columns 1-3) or that individuals who were misdiagnosed, transferred out, or whose outcome data was missing experienced unsuccessful outcomes (columns 4-6). Standard errors are provided in parentheses below each coefficient and significance levels are indicated as follows: \* $p < 0.05$ , \*\* $p < 0.01$ , \*\*\* $p < 0.001$ . Column 2 includes clinic fixed effects. Column 3 adds the following individual characteristics: language preference, sex, age, HIV status, nutritional support, presence of any comorbidities, whether the patient was infected with pulmonary or extrapulmonary TB, whether the patient was being re-treated for TB, whether the patient's diagnosis was bacteriologically confirmed, and months on treatment at the time of enrollment in the study.

**Table S.I.8:** Regressions of Unsuccessful Outcome and LTFU on Intervention Assignment, Bacteriologically Confirmed Individuals.

|  | Unsuccessful Outcome |  |  | LTFU |  |  |
| --- | --- | --- | --- | --- | --- | --- |
|  | (1) | (2) | (3) | (4) | (5) | (6) |
| Keheala | -1.3<br>(1.0) | -1.5<br>(1.0) | -1.7<br>(1.0) | -2.1**<br>(0.8) | -2.3**<br>(0.8) | -2.2*<br>(0.8) |
| Platform | -0.8<br>(1.0) | -1.2<br>(1.0) | -1.3<br>(1.0) | -1.6*<br>(0.7) | -1.9*<br>(0.8) | -1.8*<br>(0.8) |
| SMS | -1.0<br>(1.3) | -1.2<br>(1.4) | -1.2<br>(1.4) | -1.4<br>(0.9) | -1.5<br>(0.9) | -1.3<br>(0.9) |
| Clinic & Month Dummies |  | Yes | Yes |  | Yes | Yes |
| Individual Chars. |  |  | Yes |  |  | Yes |
| *N* | 11243 | 10506 | 10150 | 11243 | 10506 | 10150 |

Estimated reduction of risk (relative to the control) from linear regressions of unsuccessful outcomes (columns 1-3) and LTFU (columns 4-6) on treatment assignment, restricting to bacteriologically confirmed individuals. Standard errors are provided in parentheses below each coefficient and significance levels are indicated as follows: \* $p < 0.05$ , \*\* $p < 0.01$ , \*\*\* $p < 0.001$ . Columns 2 and 5 include clinic fixed effects. Columns 3 and 6 add the following individual characteristics: language preference, sex, age, HIV status, nutritional support, presence of any comorbidities, whether the patient was infected with pulmonary or extrapulmonary TB, whether the patient was being retreated for TB, whether the patient's diagnosis was bacteriologically confirmed, and months on treatment at the time of enrollment in the study.

**Table S.I.9:** Regressions of Unsuccessful Outcome and LTFU on Intervention Assignment, Individuals with Pulmonary TB.

|  | Unsuccessful Outcome |  |  | LTFU |  |  |
| --- | --- | --- | --- | --- | --- | --- |
|  | (1) | (2) | (3) | (4) | (5) | (6) |
| Keheala | -2.3*<br>(0.9) | -2.5**<br>(0.9) | -2.8**<br>(1.0) | -2.4***<br>(0.7) | -2.6***<br>(0.7) | -2.5***<br>(0.7) |
| Platform | -1.3<br>(0.9) | -1.5<br>(0.9) | -1.8<br>(1.0) | -1.6*<br>(0.6) | -1.8**<br>(0.7) | -1.8**<br>(0.7) |
| SMS | -1.3<br>(1.3) | -1.3<br>(1.4) | -1.4<br>(1.5) | -1.5<br>(0.9) | -1.6<br>(0.9) | -1.4<br>(0.9) |
| Clinic & Month Dummies |  | Yes | Yes |  | Yes | Yes |
| Individual Chars. |  |  | Yes |  |  | Yes |
| *N* | 13163 | 12335 | 11903 | 13163 | 12335 | 11903 |

Estimated reduction of risk (relative to the control) from linear regressions of unsuccessful outcomes (columns 1-3) and LTFU (columns 4-6) on treatment assignment, restricting to individuals with pulmonary TB. Standard errors are provided in parentheses below each coefficient and significance levels are indicated as follows: \* $p < 0.05$ , \*\* $p < 0.01$ , \*\*\* $p < 0.001$ . Columns 2 and 5 include clinic fixed effects. Columns 3 and 6 add the following individual characteristics: language preference, sex, age, HIV status, nutritional support, presence of any comorbidities, whether the patient was infected with pulmonary or extrapulmonary TB, whether the patient was being retreated for TB, whether the patient's diagnosis was bacteriologically confirmed, and months on treatment at the time of enrollment in the study.

**Table S.I.10:**
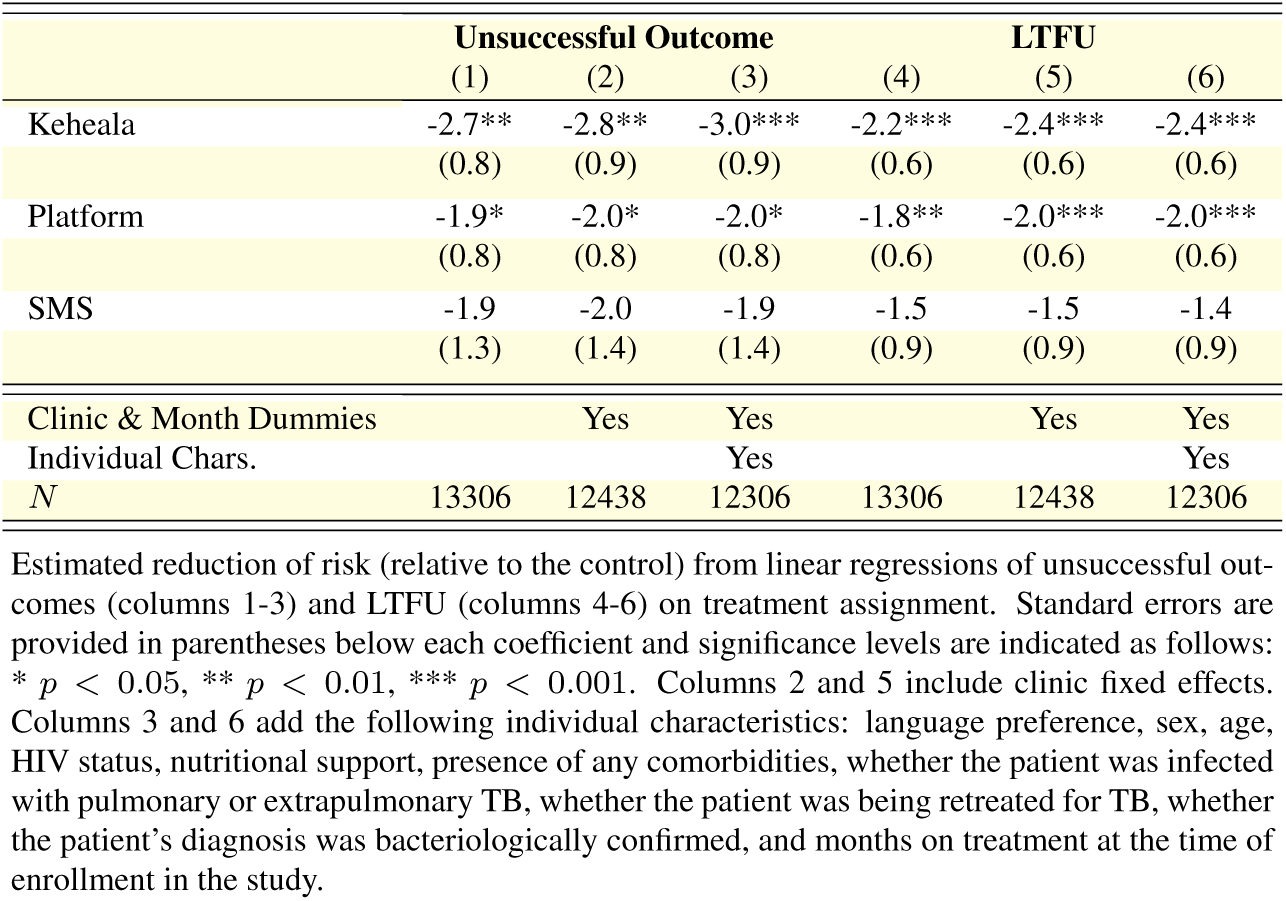
Regressions of Unsuccessful Outcome and LTFU on Intervention Assignment, Individuals Not in Retreatment.

**Table S.I.11:**
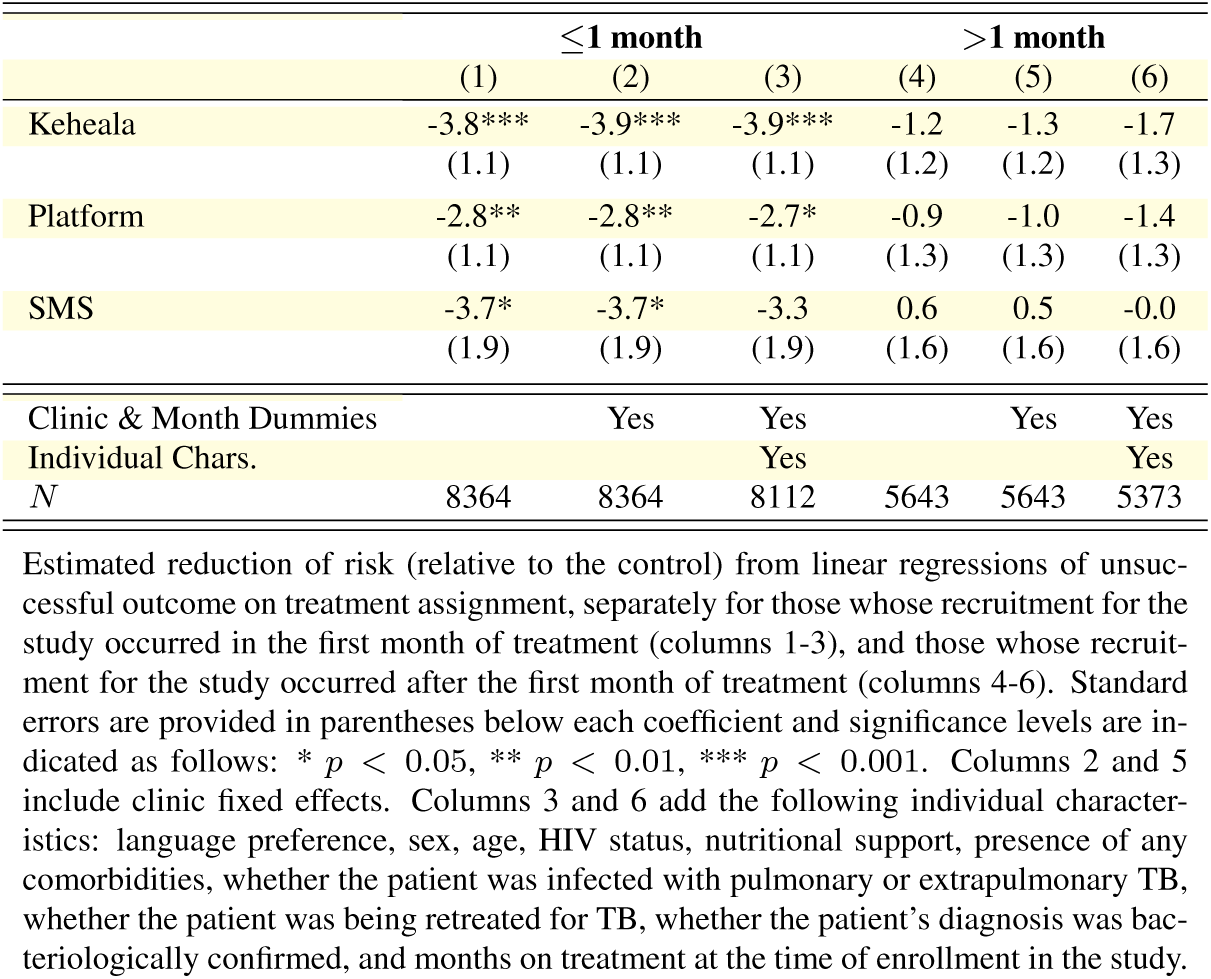
Regressions of Unsuccessful Outcome on Intervention Assignment, By Month of Treatment at Start of Intervention.

**Table S.I.12:** Regressions of LTFU on Intervention Assignment, By Month of Treatment at Start of Intervention.

| | $\leq 1$ month | | | $> 1$ month | | |
| --- | --- | --- | --- | --- | --- | --- |
|  | (1) | (2) | (3) | (4) | (5) | (6) |
| Keheala | -3.2***<br>(0.8) | -3.3***<br>(0.8) | -3.1***<br>(0.8) | -1.9*<br>(0.8) | -2.0*<br>(0.8) | -2.1*<br>(0.9) |
| Platform | -2.5**<br>(0.8) | -2.5**<br>(0.8) | -2.5**<br>(0.8) | -1.6<br>(0.8) | -1.6<br>(0.8) | -1.7<br>(0.9) |
| SMS | -2.2<br>(1.2) | -2.2<br>(1.2) | -1.9<br>(1.2) | -1.0<br>(1.1) | -1.0<br>(1.1) | -1.1<br>(1.2) |
| Clinic & Month Dummies |  | Yes | Yes |  | Yes | Yes |
| Individual Chars. |  |  | Yes |  |  | Yes |
| N | 8364 | 8364 | 8112 | 5643 | 5643 | 5373 |

Estimated reduction of risk (relative to the control) from linear regressions of LTFU on treatment assignment, separately for those whose recruitment for the study occurred in the first month of treatment (columns 1-3), and those whose recruitment for the study occurred after the first month of treatment (columns 4-6). Standard errors are provided in parentheses below each coefficient and significance levels are indicated as follows: \* $p < 0.05$ , \*\* $p < 0.01$ , \*\*\* $p < 0.001$ . Columns 2 and 5 include clinic fixed effects. Columns 3 and 6 add the following individual characteristics: language preference, sex, age, HIV status, nutritional support, presence of any comorbidities, whether the patient was infected with pulmonary or extrapulmonary TB, whether the patient was being retreated for TB, whether the patient's diagnosis was bacteriologically confirmed, and months on treatment at the time of enrollment in the study.

**Table S.I.13:** Estimated Reduction in Risk of Unsuccessful Outcome of the Keheala Intervention (Relative to the Control) Assuming Different Rates of Type I and Type II Errors in the TIBU Outcome Data.

| Type I<br>Error Rate (%) | Type II Error Rate (%) |  |  |  |  |  |
| --- | --- | --- | --- | --- | --- | --- |
|  | 0 | 1 | 2 | 3 | 4 | 5 |
| 0 | 2.61<br>(p=0.0011) | 2.59<br>(p=0.0012) | 2.56<br>(p=0.0012) | 2.53<br>(p=0.0015) | 2.51<br>(p=0.0014) | 2.48<br>(p=0.0017) |
| 2 | 2.56<br>(p=0.0012) | 2.53<br>(p=0.0015) | 2.51<br>(p=0.0014) | 2.48<br>(p=0.0017) | 2.46<br>(p=0.0016) | 2.43<br>(p=0.0019) |
| 4 | 2.51<br>(p=0.0014) | 2.48<br>(p=0.0017) | 2.46<br>(p=0.0016) | 2.43<br>(p=0.0019) | 2.40<br>(p=0.0018) | 2.38<br>(p=0.0022) |
| 6 | 2.46<br>(p=0.0016) | 2.43<br>(p=0.0019) | 2.40<br>(p=0.0018) | 2.38<br>(p=0.0022) | 2.35<br>(p=0.0020) | 2.33<br>(p=0.0025) |
| 8 | 2.40<br>(p=0.0018) | 2.38<br>(p=0.0022) | 2.35<br>(p=0.0022) | 2.33<br>(p=0.0025) | 2.30<br>(p=0.0025) | 2.27<br>(p=0.0030) |
| 10 | 2.35<br>(p=0.0022) | 2.33<br>(p=0.0025) | 2.30<br>(p=0.0025) | 2.27<br>(p=0.0030) | 2.25<br>(p=0.0027) | 2.22<br>(p=0.0033) |

Estimated reduction in risk of unsuccessful outcomes (relative to the control) of the Keheala Intervention in the presence of different rates of Type I and Type II errors. p-values from a two-sided, two-sample test of proportions are presented in parentheses below.

**Table S.I.14:**
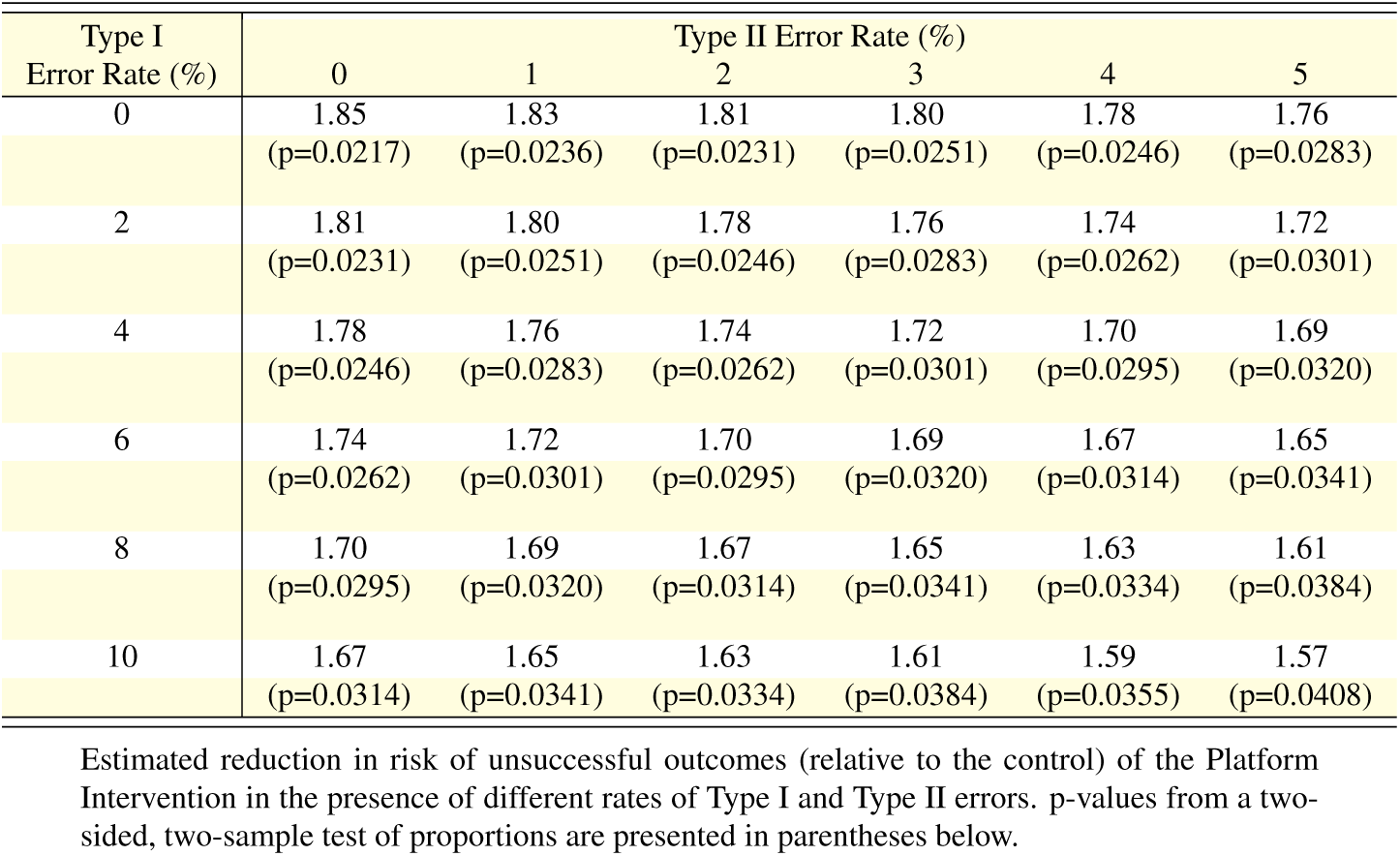
Estimated Reduction in Risk of Unsuccessful Outcome of the Platform Intervention (Relative to the Control) Assuming Different Rates of Type I and Type II Errors in the TIBU Outcome Data.

| Type I<br>Error Rate (%) | Type II Error Rate (%) |  |  |  |  |  |
| --- | --- | --- | --- | --- | --- | --- |
|  | 0 | 1 | 2 | 3 | 4 | 5 |
| 0 | 1.85<br>(p=0.0217) | 1.83<br>(p=0.0236) | 1.81<br>(p=0.0231) | 1.80<br>(p=0.0251) | 1.78<br>(p=0.0246) | 1.76<br>(p=0.0283) |
| 2 | 1.81<br>(p=0.0231) | 1.80<br>(p=0.0251) | 1.78<br>(p=0.0246) | 1.76<br>(p=0.0283) | 1.74<br>(p=0.0262) | 1.72<br>(p=0.0301) |
| 4 | 1.78<br>(p=0.0246) | 1.76<br>(p=0.0283) | 1.74<br>(p=0.0262) | 1.72<br>(p=0.0301) | 1.70<br>(p=0.0295) | 1.69<br>(p=0.0320) |
| 6 | 1.74<br>(p=0.0262) | 1.72<br>(p=0.0301) | 1.70<br>(p=0.0295) | 1.69<br>(p=0.0320) | 1.67<br>(p=0.0314) | 1.65<br>(p=0.0341) |
| 8 | 1.70<br>(p=0.0295) | 1.69<br>(p=0.0320) | 1.67<br>(p=0.0314) | 1.65<br>(p=0.0341) | 1.63<br>(p=0.0334) | 1.61<br>(p=0.0384) |
| 10 | 1.67<br>(p=0.0314) | 1.65<br>(p=0.0341) | 1.63<br>(p=0.0334) | 1.61<br>(p=0.0384) | 1.59<br>(p=0.0355) | 1.57<br>(p=0.0408) |

Estimated reduction in risk of unsuccessful outcomes (relative to the control) of the Platform Intervention in the presence of different rates of Type I and Type II errors. p-values from a two-sided, two-sample test of proportions are presented in parentheses below.

**Table S.I.15:** Estimated Reduction in Risk of Unsuccessful Outcome of the SMS Intervention (Relative to the Control) Assuming Different Rates of Type I and Type II Errors in the TIBU Outcome Data.

| Type I<br>Error Rate (%) | Type II Error Rate (%) |  |  |  |  |  |
| --- | --- | --- | --- | --- | --- | --- |
|  | 0 | 1 | 2 | 3 | 4 | 5 |
| 0 | 1.93<br>(p=0.0789) | 1.91<br>(p=0.0796) | 1.89<br>(p=0.0729) | 1.87<br>(p=0.0845) | 1.85<br>(p=0.0774) | 1.83<br>(p=0.0896) |
| 2 | 1.89<br>(p=0.0729) | 1.87<br>(p=0.0845) | 1.85<br>(p=0.0774) | 1.83<br>(p=0.0896) | 1.81<br>(p=0.0821) | 1.79<br>(p=0.0951) |
| 4 | 1.85<br>(p=0.0774) | 1.83<br>(p=0.0896) | 1.81<br>(p=0.0821) | 1.79<br>(p=0.0951) | 1.77<br>(p=0.0872) | 1.75<br>(p=0.1009) |
| 6 | 1.81<br>(p=0.0821) | 1.79<br>(p=0.0951) | 1.77<br>(p=0.0872) | 1.75<br>(p=0.1009) | 1.74<br>(p=0.0925) | 1.72<br>(p=0.1071) |
| 8 | 1.77<br>(p=0.0872) | 1.75<br>(p=0.1009) | 1.74<br>(p=0.0925) | 1.72<br>(p=0.1071) | 1.70<br>(p=0.0982) | 1.68<br>(p=0.0989) |
| 10 | 1.74<br>(p=0.0925) | 1.72<br>(p=0.1071) | 1.70<br>(p=0.0982) | 1.68<br>(p=0.0989) | 1.66<br>(p=0.1042) | 1.64<br>(p=0.1050) |

Estimated reduction in risk of unsuccessful outcomes (relative to the control) of the SMS Intervention in the presence of different rates of Type I and Type II errors. p-values from a two-sided, two-sample test of proportions are presented in parentheses below.

**Table S.I.16:** Cross-Tabulation of Treatment Outcomes Recorded in the TIBU Digital Registry and the Clinics’ Paper Registries.

| Outcome in TIBU | Treatment Outcome in Paper Registry |  |  |  |  |  | Total |
| --- | --- | --- | --- | --- | --- | --- | --- |
|  | Blank | C | D | F | LTFU | TC |  |
| Blank | 53 | 10 | 0 | 1 | 1 | 5 | 70 |
| C | 16 | 689 | 1 | 3 | 4 | 42 | 755 |
| D | 1 | 1 | 105 | 0 | 1 | 8 | 116 |
| F | 0 | 1 | 1 | 22 | 4 | 2 | 30 |
| LTFU | 0 | 2 | 0 | 0 | 95 | 4 | 101 |
| MT4 | 1 | 3 | 0 | 4 | 1 | 1 | 10 |
| NC | 9 | 28 | 6 | 0 | 4 | 30 | 78 |
| TC | 8 | 57 | 6 | 0 | 10 | 1,059 | 1,140 |
| TO | 3 | 40 | 5 | 0 | 13 | 62 | 123 |
| Total | 91 | 831 | 124 | 30 | 133 | 1,213 | 2,423 |

Individuals in a given row were all assigned the same outcome in the TIBU registry. Each column presents how many of those individuals' treatment outcomes were recorded as blank, C, D, F, *etc.* in the clinics' paper registries.

**Table S.I.17:** Type 1 and Type 2 Error Rates in the TIBU Digital Registry.

|  | Outcome in TIBU |  | Total |
| --- | --- | --- | --- |
|  | Successful | Unsuccessful |  |
| Control | 2.3 | 6.5 | 2.8 |
| SMS | 3.6 | 2.9 | 3.5 |
| Platform | 2.2 | 9.7 | 3.1 |
| Keheala | 2.6 | 7.6 | 3.1 |
| Total | 2.5 | 7.7 | 3.1 |

Error rates by unsuccessful outcome and intervention group. Errors in the 'Successful' column represent false negatives (type II errors). Errors in the 'Unsuccessful' column represent false positives (type I errors).

**Figure J.6:**
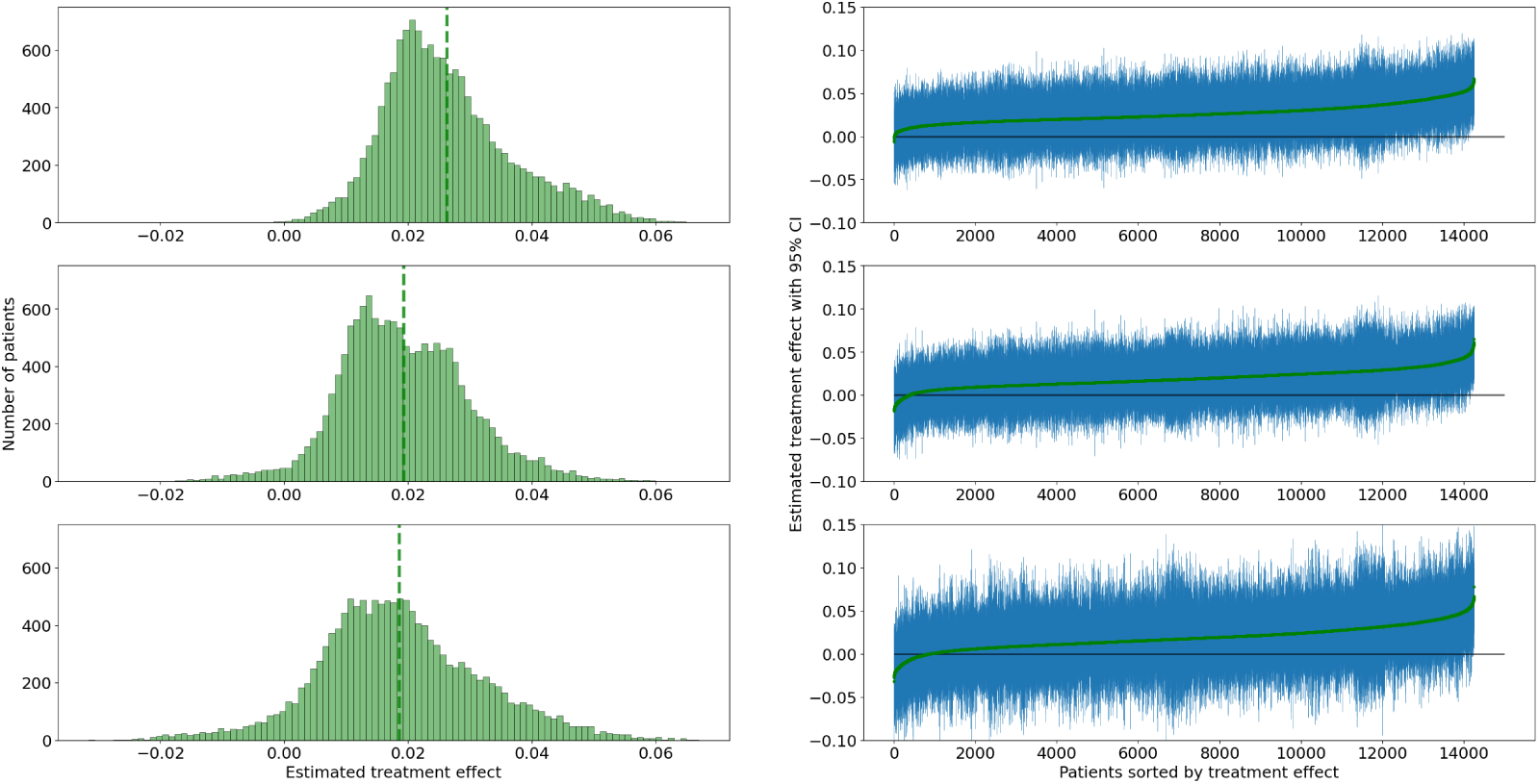
Estimated Individual Treatment Effects by Intervention The panels on the left present histograms of estimated individual effects of each intervention from a causal forest estimator (Keheala, top; platform, middle; SMS, bottom). Estimates are produced for all participants. The panels on the right present these estimates alongside confidence intervals (in blue).

## References

[1] Organization WH, et al. Gear up to end TB: introducing the end TB strategy. World Health Organization; 2015.

[2] Reid MJ, Arinaminpathy N, Bloom A, Bloom BR, Boehme C, Chaisson R, et al. Building a tuberculosis-free world: The Lancet Commission on tuberculosis. The Lancet. 2019;393(10178):1331–84.

[3] Subbaraman R, de Mondesert L, Musiimenta A, Pai M, Mayer KH, Thomas BE, et al. Digital adherence technologies for the management of tuberculosis therapy: mapping the landscape and research priorities. BMJ global health. 2018;3(5):e001018.

[4] Ngwatu BK, Nsengiyumva NP, Oxlade O, Mappin-Kasirer B, Nguyen NL, Jaramillo E, et al. The impact of digital health technologies on tuberculosis treatment: a systematic review. European Respiratory Journal. 2018;51(1).

[5] Subbaraman R, Jhaveri T, Nathavitharana RR. Closing gaps in the tuberculosis care cascade: an action-oriented research agenda. Journal of clinical tuberculosis and other mycobacterial diseases. 2020;19:100144.

[6] Organization WH. WHO Consolidated Guidelines on Tuberculosis, Module 4: Treatment - Drug-Resistant Tuberculosis Treatment; 2022. Last accessed July 11, 2022 at https://www.who.int/publications/i/item/9789240047716.

[7] Mohamed MS, Zary M, Kafie C, Chilala CI, Bahukudumbi S, Foster N, et al. The impact of digital adherence technologies on treatment outcomes, adherence, and patient-reported outcomes in tuberculosis: a systematic review and meta-analysis. BMC Infectious Diseases. 2025;25(1):1314.

[8] Rand DG, Yoeli E, Hoffman M. Harnessing reciprocity to promote cooperation and the provisioning of public goods. Policy Insights from the Behavioral and Brain Sciences. 2014;1(1):263–9.

[9] Yoeli E, Rathauser J, Bhanot SP, Kimenye MK, Mailu E, Masini E, et al. Digital health support in treatment for tuberculosis. New England Journal of Medicine. 2019;381(10):986–7.

[10] Organization WH. Global tuberculosis report 2020. World Health Organization; 2020.

[11] Kenya Tuberculosis Treatment Success Rate, 2022; 2025. In 2022, Kenya’s tuberculosis treatment success rate was 89.0%, implying 11.0% treatment non-completion. CDC Kenya HIV & TB Overview; World Bank Development Indicators. Available from: https://www.cdc.gov/global-hiv-tb/php/where-we-work/kenya.html `and` https://data.worldbank.org/indicator/SH.TBS.CURE.ZS.

[12] Kariuki J. Kenyan mobile phone users up to 38 million; 2016. Last accessed May 11, 2018 at https://www.nation.co.ke/ business/Kenyan-mobile-phone-users-38-million/996-3023970-13ydsf4z/index.html.

[13] Communications Authority of Kenya. First quarter sector statistics report for the financial year 2015/2016; 2016. Last accessed May 11, 2018 at http://www.ca.go.ke/index.php/what-we-do/94-news/366-kenya-s-mobile-penetration-hits-88-per-cent.

[14] Kemibaro M. Kenya’s latest 2016 mobile and Internet statistics; 2016. Last accessed May 11, 2018 at http://www.moseskemibaro.com/2016/10/01/kenyas-latest-2016-mobile-internet-statistics/.

[15] National Tuberculosis, Leprosy, and Lung Disease Program. Guidelines for TB Infection Prevention and Control for Health Care workers in Kenya 2021; 2021. Last accessed January 18, 2022 at https://www.nltp.co.ke/wp-content/uploads/2022/01/IPC_-Guidelines_-2021_Final_14_06_2021.pdf.

[16] National Tuberculosis, Leprosy, and Lung Disease Program. Guidelines for Management of Tuberculosis and Leprosy in Kenya; 2013. Last accessed January 18, 2022 at https://nltp.co.ke/wp-content/uploads/2020/10/TB_Treatment_GUIDELINES_2013.pdf.

[17] Massachusetts Institute of Technology. Behavior Change and Digital Health Interventions for Improved TB Treatment Outcomes; 2019. Available for download at https://clinicaltrials.gov/ct2/show/NCT04119375.

[18] Kraft-Todd G, Yoeli E, Bhanot S, Rand D. Promoting cooperation in the field. Current Opinion in Behavioral Sciences. 2015;3:96–101.

[19] Thomas BE, Kumar JV, Chiranjeevi M, Shah D, Khandewale A, Thiruvengadam K, et al. Evaluation of the accuracy of 99DOTS, a novel cellphone-based strategy for monitoring adherence to tuberculosis medications: comparison of DigitalAdherence data with urine isoniazid testing. Clinical Infectious Diseases. 2020;71(9):e513–6.

[20] Subbaraman R, Thomas BE, Kumar JV, Lubeck-Schricker M, Khandewale A, Thies W, et al. Measuring tuberculosis medication adherence: a comparison of multiple approaches in relation to urine isoniazid metabolite testing within a cohort study in India. In: Open Forum Infectious Diseases. vol. 8. Oxford University Press US; 2021. p. ofab532.

[21] Liu X, Lewis JJ, Zhang H, Lu W, Zhang S, Zheng G, et al. Effectiveness of electronic reminders to improve medication adherence in tuberculosis patients: a cluster-randomised trial. PLoS medicine. 2015;12(9):e1001876.

[22] Imperial MZ, Nahid P, Phillips PP, Davies GR, Fielding K, Hanna D, et al. A patient-level pooled analysis of treatment-shortening regimens for drug-susceptible pulmonary tuberculosis. Nature medicine. 2018;24(11):1708–15.

[23] Collins D, Njuguna C. The economic cost of non-adherence to TB medicines resulting from stock-outs and loss to follow-up in Kenya. Submitted to the US Agency for international development by the Systems for Improved Access to pharmaceuticals and services (SIAPS) program Arlington: Management Sciences for Health. 2016.

[24] Ochalek J, Lomas J, Claxton K. Estimating health opportunity costs in low-income and middle-income countries: a novel approach and evidence from cross-country data. BMJ global health. 2018;3(6):e000964.

[25] Mohammed S, Glennerster R, Khan AJ. Impact of a daily SMS medication reminder system on tuberculosis treatment outcomes: a randomized controlled trial. PloS one. 2016;11(11):e0162944.

[26] Park S, Sentissi I, Gil SJ, Park WS, Oh B, Son AR, et al. Medication event monitoring system for infectious tuberculosis treatment in Morocco: a retrospective cohort study. International journal of environmental research and public health. 2019;16(3):412.

[27] Story A, Aldridge RW, Smith CM, Garber E, Hall J, Ferenando G, et al. Smartphone-enabled video-observed versus directly observed treatment for tuberculosis: a multicentre, analyst-blinded, randomised, controlled superiority trial. The Lancet. 2019;393(10177):1216–24.

[28] Wang N, Shewade HD, Thekkur P, Zhang H, Yuan Y, Wang X, et al. Do electronic medication monitors improve tuberculosis treatment outcomes? programmatic experience from China. PLoS One. 2020;15(11):e0242112.

[29] Tadesse AW, Mohammed Z, Foster N, Quaife M, McQuaid CF, Levy J, et al. Evaluation of implementation and effectiveness of digital adherence technology with differentiated care to support tuberculosis treatment adherence and improve treatment outcomes in Ethiopia: a study protocol for a cluster randomised trial. BMC infectious diseases. 2021;21(1):1–11.

[30] Sahile Z, Perimal-Lewis L, Arbon P, Maeder AJ. Protocol of a parallel group Randomized Control Trial (RCT) for Mobile-assisted Medication Adherence Support (Ma-MAS) intervention among Tuberculosis patients. PloS one. 2021;16(12):e0261758.

[31] Sekandi JN, Onuoha NA, Buregyeya E, Zalwango S, Kaggwa PE, Nakkonde D, et al. Using a Mobile Health Intervention (DOT Selfie) With Transfer of Social Bundle Incentives to Increase Treatment Adherence in Tuberculosis Patients in Uganda: Protocol for a Randomized Controlled Trial. JMIR Research Protocols. 2021;10(1):e18029.

[32] Cattamanchi A, Crowder R, Kityamuwesi A, Kiwanuka N, Lamunu M, Namale C, et al. Digital adherence technology for tuberculosis treatment supervision: A stepped-wedge cluster-randomized trial in Uganda. PLoS medicine. 2021;18(5):e1003628.

[33] Acosta J, Flores P, Alarcón M, Grande-Ortiz M, Moreno-Exebio L, Puyen Z. A randomised controlled trial to evaluate a medication monitoring system for TB treatment. The International Journal of Tuberculosis and Lung Disease. 2022;26(1):44–9.

[34] Marx F, Meehan S, Jivan D, Dunbar R, Hoddinott G, Hesseling A, et al. Use of interactive messaging to reduce pre-diagnosis loss to follow-up for TB care. The International Journal of Tuberculosis and Lung Disease. 2022;26(1):26–32.

[35] Alwood K, Keruly J, Moore-Rice K, Stanton DL, Chaulk CP, Chaisson RE. Effectiveness of supervised, intermittent therapy for tuberculosis in HIV-infected patients. AIDS (London, England). 1994;8(8):1103–8.

[36] Veinot TC, Mitchell H, Ancker JS. Good intentions are not enough: how informatics interventions can worsen inequality. Journal of the American Medical Informatics Association. 2018;25(8):1080–8.

[37] Boutilier JJ, Jónasson JO, Yoeli E. Improving Tuberculosis Treatment Adherence Support: The Case for Targeted Behavioral Interventions. Manufacturing & Service Operations Management. 2021.

[38] Yoeli E, Hoffman M, Rand DG, Nowak MA. Powering up with indirect reciprocity in a large-scale field experiment. Proceedings of the National Academy of Sciences. 2013;110(Supplement 2):10424–9.

[39] Athey S, Tibshirani J, Wager S. Generalized random forests. The Annals of Statistics. 2019;47(2):1148–78.

[40] Tibshirani J, Athey S, Sverdrup E, Wager S. Generalized Random Forests; 2017. Last accessed January 28, 2022 at https://grf-labs.github.io/grf/authors.html.

